# SARS-CoV-2 testing strategies to contain school-associated transmission: model-based analysis of impact and cost of diagnostic testing, screening, and surveillance

**DOI:** 10.1101/2021.05.12.21257131

**Authors:** Alyssa Bilinski, Andrea Ciaranello, Meagan C. Fitzpatrick, John Giardina, Maunank Shah, Joshua A. Salomon, Emily A. Kendall

## Abstract

**Background:** In March 2021, the Biden administration allocated $10 billion for COVID-19 testing in schools. We evaluate the costs and benefits of testing strategies to reduce the infection risks of full-time in-person K-8 education at different levels of community incidence.

**Methods:** We used an agent-based network model to simulate transmission in elementary and middle school communities, parameterized to a US school structure and assuming dominance of the delta COVID-19 variant. We assess the value of different strategies for testing students and faculty/staff, including expanded diagnostic testing (“test to stay” policies that take the place of isolation for symptomatic students or quarantine for exposed classrooms); screening (routinely testing asymptomatic individuals to identify infections and contain transmission); and surveillance (testing a random sample of students to signaling undetected transmission and trigger additional investigation or interventions).

**Main outcome measures:** We project 30-day cumulative incidence of SARS-CoV-2 infection; proportion of cases detected; proportion of planned and unplanned days out of school; and the cost of testing programs and of childcare costs associated with different strategies. For screening policies, we further estimate cost per SARS-CoV-2 infection averted in students and staff, and for surveillance, probability of correctly or falsely triggering an outbreak response at different incidence and attack rates.

**Results:** Accounting for programmatic and childcare costs, “test to stay” policies achieve similar model-projected transmission to quarantine policies, with reduced overall costs. Weekly universal screening prevents approximately 50% of in-school transmission, with a lower projected societal cost than hybrid or remote schooling. The cost per infection averted in students and staff by weekly screening is lower for older students and schools with higher mitigation and declines as community transmission rises. In settings where local student incidence is unknown or rapidly changing, surveillance may trigger detection of moderate-to-large in-school outbreaks with fewer resources compared to screening.

**Conclusions:** “Test to stay” policies and/or screening tests can facilitate consistent in-person school attendance with low transmission risk across a range of community incidence. Surveillance may be a useful reduced-cost option for detecting outbreaks and identifying school environments that may benefit from increased mitigation.

## Introduction

In K-12 education, COVID-19 poses risks to student and teacher health, school operations, and local communities. As of May 2021, about a third of US students were not offered the option of full-time in-person attendance and, where in-person schooling was offered, a substantial proportion of families opted for remote learning (1–3). However, virtual and hybrid models imposed substantial burdens, including educational and mental health risks for students, training and logistical challenges for teachers, and productivity or child-care costs for the working parents of younger students (4–10). Districts are currently making plans for the fall with a priority on safe in-person education, even in the event of seasonal increases in transmission and reduced vaccine efficacy against new variants (11–16).

In March 2021, the Biden administration allocated $10 billion for diagnostic and screening tests in schools (17). Improvements in SARS-CoV-2 diagnostic technology and infrastructure – such as increased PCR testing capacity, lateral flow rapid antigen tests, and validations of specimen pooling – make frequent, widespread testing a viable option (18,19). A key question is how to best allocate this funding to maximize in-person educational time while both controlling COVID-19 transmission and managing financial and operational costs. Centers for Disease Control and Prevention (CDC) guidelines for school reopening divide testing into three categories (20). Diagnostic testing targets those showing symptoms of COVID-19 as well as close contacts of a diagnosed case. Screening entails routine asymptomatic testing of the full school population in order to identify active cases and prevent onward transmission. Last, surveillance testing involves sampling a fraction of the population in order to identify potential outbreaks and trigger additional public health responses (e.g., school-wide screening or school closures). At present, schools require guidance on how to best allocate resources toward different testing objectives.

In this manuscript, we address several questions regarding the role of testing in educational settings: First, to what extent can different testing strategies limit school-associated transmission of SARS-CoV-2 while sustaining in-person learning? How frequent are quarantines arising from different strategies, and to what extent can testing of contacts avert days out of school? How do testing costs compare to the financial costs associated with school absences or closures? Last, how might these outcomes vary depending on local transmission risk? We focus on elementary and middle schools, both because childcare costs are more substantial for these groups and because vaccines are not yet authorized for all students of these ages (21). We use an agent-based simulation of COVID-19 transmission to compare outcomes associated with different testing strategies, with a particular focus on infections, in-person educational days, and costs.

## Methods

We used a previously validated agent-based simulation model to estimate the effects of different testing strategies in elementary and middle schools in the US (Figure S1) (22). The model incorporates interactions between individuals in school, household, and out-of-school childcare settings, as well as infections introduced exogenously through other community interactions.

### Model structure

For a simulated elementary school (638 students in grades K-5 and 60 staff) and middle school (460 students in grades 6-8 and 51 staff), we generated households from synthetic population data (23) and grouped students into fixed classroom cohorts with a primary teacher. In schools, individuals had sustained daily contact with their classroom cohort as well as additional interactions with other members of the school community. Outside of schools, in addition to an exogenous community infection risk, individuals interacted with household members, and each day that students did not attend school, families mixed with another randomly chosen family from the same school to reflect “learning pods,” social interactions, or shared childcare.

We assumed that elementary school students are half as susceptible and half as infectious as adults and that middle school students have similar susceptibility and infectiousness as adults (22,24–28). In the base case, we assumed schools adopted high mitigation (i.e., masking, ventilation, and distancing) and calibrated the model assuming the delta variant is approximately twice as transmissible as wild type (29,30). We further assumed that 90% of teachers and staff and 50% of middle school students were vaccinated with an 80% efficacious vaccine (31,32). Further details of model structure, assumptions, and validation are described in (22) and the Supplement.

### Testing strategies

#### Scenarios without testing

We first modeled three scenarios without school-based testing: 1) five-day in-person attendance (the base case, and also the schedule assumed for all testing scenarios), 2) a hybrid model in which half of each class attends school on Monday and Tuesday and the other half on Thursday and Friday (a strategy used in 2020-21) and 3) fully remote learning (a proxy for anticipated infection risk unrelated to in-person education). In these scenarios, we assumed that individuals with clinically-identifiable symptoms isolated and underwent testing outside of school on the day that symptoms appeared, that they received results within 48 hours, and that the entire classroom cohort of a diagnosed COVID-19 case was quarantined for 10 days (26). We assumed that isolation and/or diagnostic testing for symptoms caused by non-COVID etiologies occurred in 1% of students and staff each week (33).

#### Diagnostic testing

The “test to stay” strategy altered both how symptomatic students/staff are managed and how the school responds to diagnosed COVID-19 cases (34). Individuals with clinically identifiable symptoms remained at school and took a rapid test each day they had symptoms, isolating only after testing positive. Likewise, after exposure to a confirmed case, contacts remained in school and received a rapid test daily for one week (34). We assumed 80% test sensitivity during the infectious period and 100% specificity, following a second confirmatory test (35,36). We present both quarantine and test to stay for each of the five-day in-person scenarios modeled.

#### Screening and surveillance

Screening entailed weekly PCR screening (on Mondays) of all students and teachers, with 90% coverage, 90% sensitivity during infectiousness, and a 24-hour test turnaround time. Surveillance entailed weekly PCR testing (90% sensitivity) of 10-20% of the school population, randomly selected solely from unvaccinated individuals. Due to the small proportion of the school tested, if ≥1 case was detected during surveillance, 90% of the school was screened the following day, including vaccinated individuals. If further cases were found, the school changed to weekly school-wide (90% coverage) screening beginning the next Monday for the remainder of the month; otherwise, surveillance resumed as scheduled the following Monday. (Threshold selection is discussed further in the Supplement.)

Based on recent CDC guidance (16), we assumed that vaccinated individuals do not quarantine and are not included in surveillance. However, due to reduced vaccine efficacy with the delta variant and associated recommendations to test vaccinated contacts (16), we included them in “test to stay” measures and school-wide screening.

### Costs

We based screening and surveillance costs on pooled PCR testing. We assumed separate anterior nasal swab specimens were collected from each person tested, samples from up to 8 specimens were pooled and run as a single PCR, and residual individual specimens were held for immediate testing when the corresponding pool was positive (Supplement). Costs of PCR testing were estimated at $40 per assay, plus an $8 per-person cost of labor and supplies for nasal swab collection (Table 1). Rapid testing for the “test to stay” scenario cost $6 per assay plus the same specimen collection costs as for PCR specimens. In a sensitivity analysis, we also considered rapid testing with confirmatory PCR for screening and surveillance. Costs of non-school-based diagnostic testing were excluded in order to focus on the tests costs incurred by the school; this exclusion will result in conservative estimates of the societal cost savings of the “test to stay” strategy.

**Table 1:** Model parameters.

|  | Estimate | Sources/Notes |
| --- | --- | --- |
| **Key transmission model parameters (see (22) for full list and sources)** |  |  |
| Duration of infectiousness | Lognormal (5, 2) | Calibrated to match serial interval (46,47).<br>This ensures that early high transmissibility is captured, though a long tail of reduced infectiousness likely exists. (See sensitivity analyses.) |
| Classroom adult-adult symptomatic daily attack rate | 2%<br>(1% or 4% in sensitivity analysis) | Daily transmission rate between two unvaccinated adults during shared full-day contact<br>The model further adjusts for reduced elementary school students susceptibility (RR=0.5) + infectiousness (RR=0.5); asymptomatic middle school students + adults (RR=0.5).<br>See (22) for details |
| Relative attack rate for random school contacts (vs. classroom) | 0.13 | Based on 45 minutes/day of exposure |
| Household attack rate | 20% | (48,49) |
| Probability of fully asymptomatic disease | 20%, children (elementary + high school)<br>40%, adults | (24,25,28,50) |
| Probability that disease has clinically recognizable symptoms | 20%, children (elementary + high school)<br>40%, adults | (50,51) |
| Presymptomatic period (days) | Normal<br>(1.2, 0.4) | (52) |
| School size | Elementary: 638 students, 60 teachers/staff, 30 classes<br>Middle: 460 students, 51 teachers/staff, 21 classes | (53) |
| Community COVID-19 notification rate | Varied between 1 and 100 diagnosed cases per 100,00 population per day |  |
| Case detection ratio in community | 1/3 | Older US modeling estimate and current UK surveillance estimate (54,55)<br>There is some evidence that this may be low in the current wave of infections; surveillance or screening can help to ascertain the true value. |
| Vaccine effectiveness | 80% | (31,32) |
| Teacher vaccination uptake | 90% | (56), assuming full completion of regimens among those who received their first dose by April + 10% additional uptake |
| **Testing parameters** |  |  |
| **PCR** |  |  |
| Sensitivity of PCR testing during infectious period for screening + surveillance | 0.9 | (57–60). Combined with 90% screening uptake, 81% of infectious students and staff are detected. |
| Frequency of testing | 0, 1x, or 2x per week | Testing is assumed to occur on Monday +- Thursday |
| School-based screening test turnaround time | 1 day |  |
| Time from symptom onset to result of community-based diagnostic tests | 2 days |  |
| Duration of isolation after COVID-19 diagnosis | 10 days | (61) |
| Duration of quarantine after COVID-19 exposure | 10 days | (61) |
| **Rapid testing** |  |  |
| Sensitivity of rapid test during infectious period for test-to-stay | 0.8 | Estimates of culturable infections from (35) for asymptomatic individuals; for symptomatic children from (62); see further discussion in (36) |
| School-based test-to-stay turnaround time | 15 minutes (same-day isolation of positive cases) |  |
| **Costs** |  |  |
| Cost per PCR run (per 8-sample pool, and per individual in pool for | \$40 | Consistent with prices paid by early adopters (63), Massachusetts school testing, and some types of Medicare reimbursement (64) |
| testing after a positive pooled result) |  |  |
| Cost per rapid test run | \$6 | Assumes 50% discount from retail prices per documented bulk rates (65), consistent with other analyses (66) |
| Added cost per specimen collected (both PCR and rapid) | \$8 | (67) |
| Cost per planned day at home | \$35.50 | Based on group childcare costs for pre-kindergarten (68); summertime childcare costs for school-aged children are similar (69) |
| Cost per unplanned day at home | \$85.90 | Based on childcare worker wages (37) |

In comparing the costs associated with remote learning to the costs of testing, we took a modified societal perspective that focused on childcare or parent productivity costs; educational and other student costs are likely to accrue, but difficult to estimate. For remote and hybrid education, we estimated the cost of a planned day of remote instruction based on the average cost of group childcare (Table 1). For unplanned days at home (i.e., while isolated due to COVID-19 diagnosis or symptoms, or quarantined due to COVID-19 exposure), we estimated costs based on the average child care worker’s wages over a 7-hour day (37) to account for the higher costs of last-minute scheduling or inability to use group childcare (Table 1). While parents may choose to supervise remote learning at home, we assumed that the average productivity loss of supervising at-home learning was comparable to childcare costs.

### Outcomes

For each scenario, we ran the model 1000 times for 30 days each, and estimated the following outcomes over a 30-day period: average cumulative true incidence of SARS-CoV-2 infection among staff and students (not counting secondary transmissions to household members or community contacts), cumulative case detection (as a proportion of all students and staff and as an absolute number with confirmed infection during the month), detection fraction (the ratio between cases detected and true infections), and proportion of weekdays spent at home (for “unplanned” isolation or quarantine reasons, or for “planned” days at home dictated by the virtual/hybrid schedule). We performed one-way sensitivity analyses for multiple parameters to evaluate uncertainty in the number of infections (among students and teachers) prevented by weekly screening. The model was implemented in R 4.0.2. Model code is publicly available as an R package at: https://github.com/abilinski/BackToSchool2.

## Results

### Effect of in-person school attendance on COVID-19 incidence

Figure 1 and Table S1 show 30-day incidence, case detection, and school attendance outcomes of different testing scenarios. At the elementary school level, compared to fully remote instruction, 5-day in-person attendance with no in-school testing was associated with a 40% projected increase in COVID incidence among students (mean 1.9 additional infections per school per month) at a community notification rate of 10/100k/day and a 38% increase (8 additional infections per school per month) at 50 community notifications/100k/day (Figure 1A). If students with known exposures were allowed to stay in school with daily testing (the “test to stay” strategy), slightly more transmission occurred; e.g., a 43% increase over the remote-instruction baseline, at 10 community notifications/100k/day.

**Figure 1:**
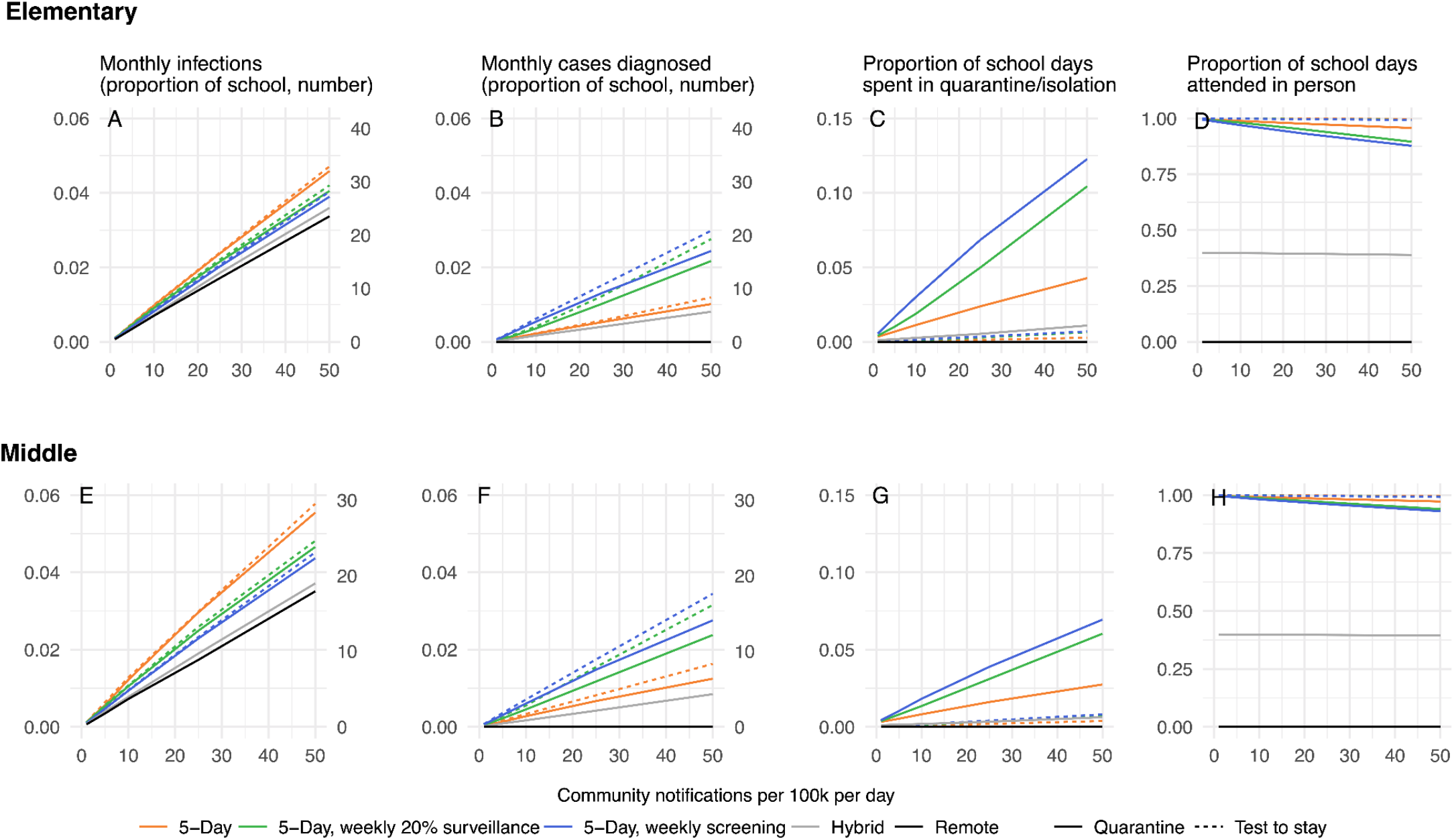
One-month cumulative incidence, case detection, isolation/quarantine, and remote learning days with multiple school schedules and testing frequencies. Results are shown over a range of community COVID-19 notification rates for an elementary school of 638 students and a middle school of 460 students. Infections (panels A and E) and diagnoses (panels B and F) are shown both as a proportion of all students and staff infected or proportion with detected cases per month (left-hand y axes) and as an expected number of infections/diagnoses among students and staff per school per month (right-hand y axes); these outcomes do not include infections among others in the community that may result from school-associated transmission. Panels C and G show the average proportion of weekdays that students and staff were scheduled to attend school but are in isolation or quarantine due to COVID-19 symptoms, diagnosis, or exposure. Panels D and H show the proportion of weekdays that student and staff attend in person after accounting for the scheduling model and isolation/quarantine. The detection fraction as reported in the text reflects the absolute number of diagnosed cases (panels B and F) divided by true cumulative incidence (panels A and E).

In the middle school where students were more susceptible and more infectious, in-person attendance had greater potential to increase transmission, although 50% student vaccination kept it partially in check. Compared to remote instruction, 5-day middle school attendance (with quarantine of known close contacts) increased incidence by 72% (3 added infections per school per month) at a community notification rate of 10/100k/day and by 60% (10 added infections per school per month) at 50 community notifications/100k/day (Figure 1E). As in the elementary school, the “test to stay” strategy increased transmission slightly compared to the remote-only baseline; e.g. from a 72% increase with quarantine to an 82% increase with test-to-stay at 10 community notifications/100k/day.

In comparison, a hybrid (A/B) schedule could prevent much of this excess transmission by reducing the number and duration of contacts, but with the downside of ≥60% remote instruction (Figure 1, gray lines).

### Transmission impact of weekly screening and surveillance

Weekly screening of all students and teachers, with isolation of the identified cases and quarantine of their unvaccinated classroom contacts, eliminated a large proportion of the incremental transmission associated with school attendance. In a community with 10 notifications/100k/day, weekly screening averted 57% of excess incidence relative to remote learning in both the elementary and the middle school. The number of infections that screening could prevent among students and teachers increased roughly in proportion to the community incidence (Figure 1 A and E, and Table S1).

At low community incidence, weekly surveillance (focusing on the unvaccinated, and converting to school-wide testing if a case is detected in surveillance) achieved a large transmission benefit relative to the number of students undergoing surveillance (Figure 1 A and E green lines, and Table 1S). For a middle school in a community with 10 notifications/100k/day, weekly 20% surveillance averted 34% of the excess transmission associated with school attendance, more than half of the 57% averted by weekly universal screening of 90% of students and educators/staff; a switch from 20% surveillance to screening the entire school was required in fewer than one third of the simulations while maintaining only 20% surveillance for the full month in more than two thirds of simulations (Figure 1 and Figure S2). At high levels of community incidence, surveillance achieved nearly the same transmission benefit as universal screening, but this impact was attributable to a high probability of detecting multiple cases and therefore converting to universal screening (reaching 99.9% probability at 100 community notifications/100k/day) (Figure S2, left panel). Across all scenarios, the probability of no in-school transmission when screening was triggered was below 25%, decreasing with increased community incidence (Figure S2, middle panels).

As in the no-screening scenario, a “test to stay” strategy after case detection slightly diminished the transmission benefits of screening or surveillance. As an example, with “test to stay” instead of quarantine for an elementary school and 10 community notifications/100k/day, weekly screening prevented 46% rather than 57% of excess transmission, and weekly 20% surveillance prevented 17% rather than 25% (Figure 1 A, comparing dashed and solid lines).

### Case detection and in-person learning days lost from screening and surveillance

At the elementary level, when the community notification rate was 10 cases/100k/day, weekly screening increased the detection fraction from 23% to 66%, and weekly 20% surveillance increased it from 23% to 39%. Despite the corresponding reduction in the absolute number of infections, the number of absolute cases detected increased (by more than a factor of two, for weekly screening), and the days spent in isolation or quarantine increased by a similar factor (Figure 1B, 1C, 1F, 1G). In an elementary school with weekly screening, the result was an average of 0.6 quarantine/isolation days per student per month at 10 community notifications/100k/day and 2.6 quarantine/isolation days per student per month at 50 community notifications/100k/day (Figure 1C). In middle school, quarantine of only unvaccinated students more than offsets the higher transmission, resulting in slightly fewer isolation or quarantine days per student than in the elementary school (Figure 1F-G).

A “test to stay” strategy had the benefit of far fewer days spent in isolation and quarantine, averaging <0.2 days per student per month, even at the highest modeled rates of community transmission and paired with maximal case detection through weekly screening.

### Costs

Testing and childcare costs over the 30-day period were estimated for each strategy (Figure 2, breakdowns in S3-S6). When no pooled screening tests were positive and all students and staff were in attendance, weekly screening cost approximately $69 per student per month (requiring, for example, 3,141 specimen collections and 465 PCR tests per month in the elementary school). At higher incidence, the testing costs of weekly screening remained roughly stable (with the increasing cost of deconvoluting positive pools partially offset by the reduced screening days due to quarantine); costs of surveillance approached those of screening as positive surveillance tests and subsequent conversion to weekly screening become common above community notification rates of 10 to 25/day (Figures S3-S4).

**Figure 2:**
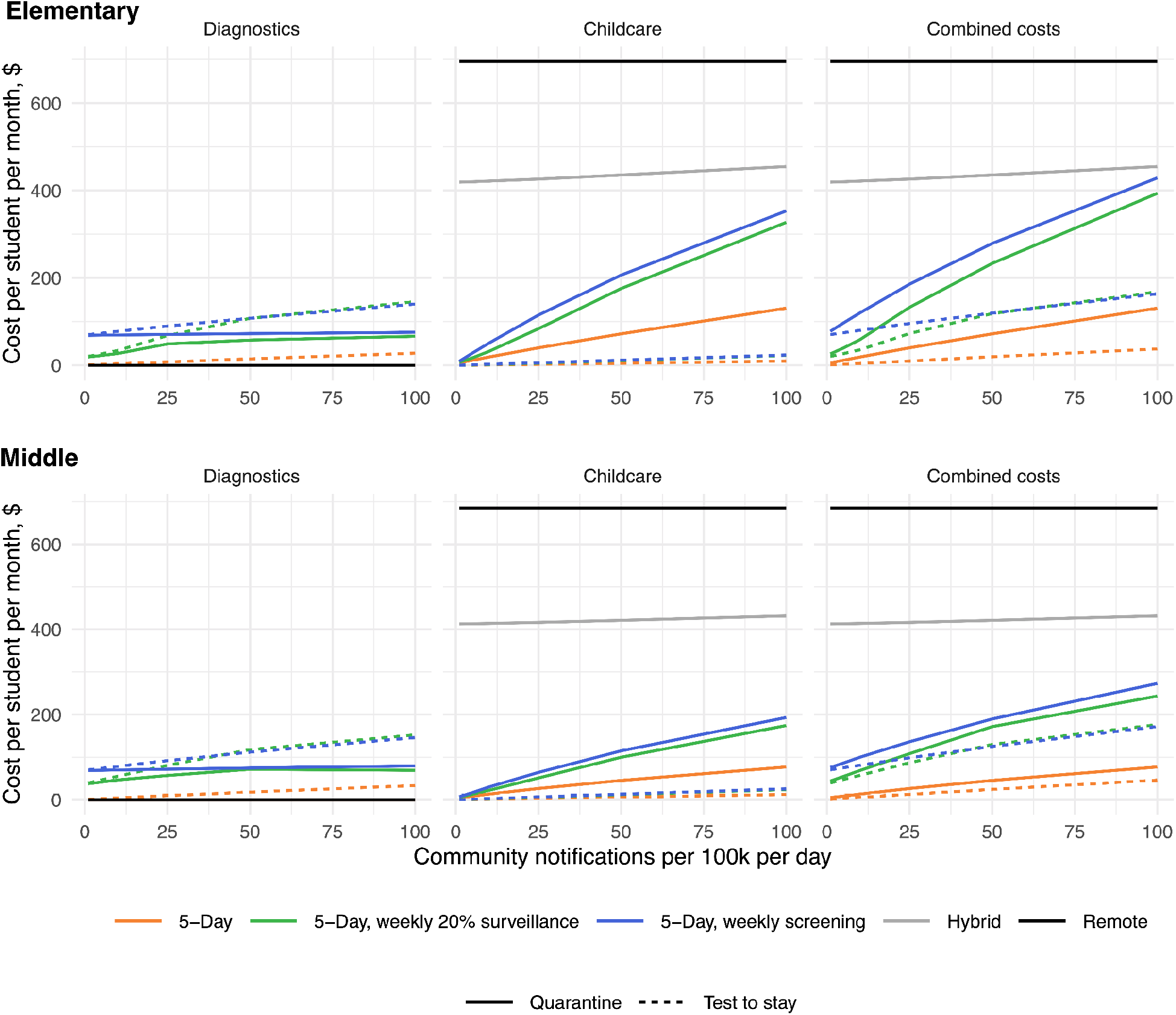
Costs associated with in-school COVID-19 testing and/or out-of-school childcare for different risk-reduction strategies, at varying community notification rates.

Accounting for childcare during isolation and quarantine, the societal costs associated with weekly screening in an elementary school ranged from <$100 per student per month at community notification rates ≤5/100k/day, to $429/student/month at a community notification rate of 100/100k/day (Figure 2, top-right panel). A “test to stay” strategy increased testing costs by an amount proportional to the community incidence (e.g., by 27% at a community notification rate of 25/100k/day), but reduced the combined costs of testing plus child care at all community notification rates (Figure 2, dotted lines). In comparison, the cost of childcare exceeded $400/student/month for a hybrid schedule and exceeded $600/student/month for a fully-remote schedule at all incidence levels. The estimated costs of a rapid antigen screening strategy were similar to those of pooled PCR screening (Figure S7).

### Cost per infection averted

We estimated the cost of screening or surveillance per infection averted among students and teachers/staff, when compared to the same full-time in-person attendance without school-based testing (Figure 3). In the elementary school, combined costs per infection directly averted were between $4,000 and $20,000 (depending on the community incidence and the use of quarantine or test-to-stay) at community notification rates of ≥25 cases/100k/day; costs rose to approximately $50,000 per infection averted at 10 cases/100k/day and $500,000 per infection averted at 1 case/100k/day. In a middle school with a strategy of screening and quarantine, the greater risk of transmission reduces the costs of per infection averted by approximately half compared to the elementary school, despite the comparative inefficiency of screening vaccinated students: for example, $25,000 at 10 cases/100k/day (Figure 3).

**Figure 3:**
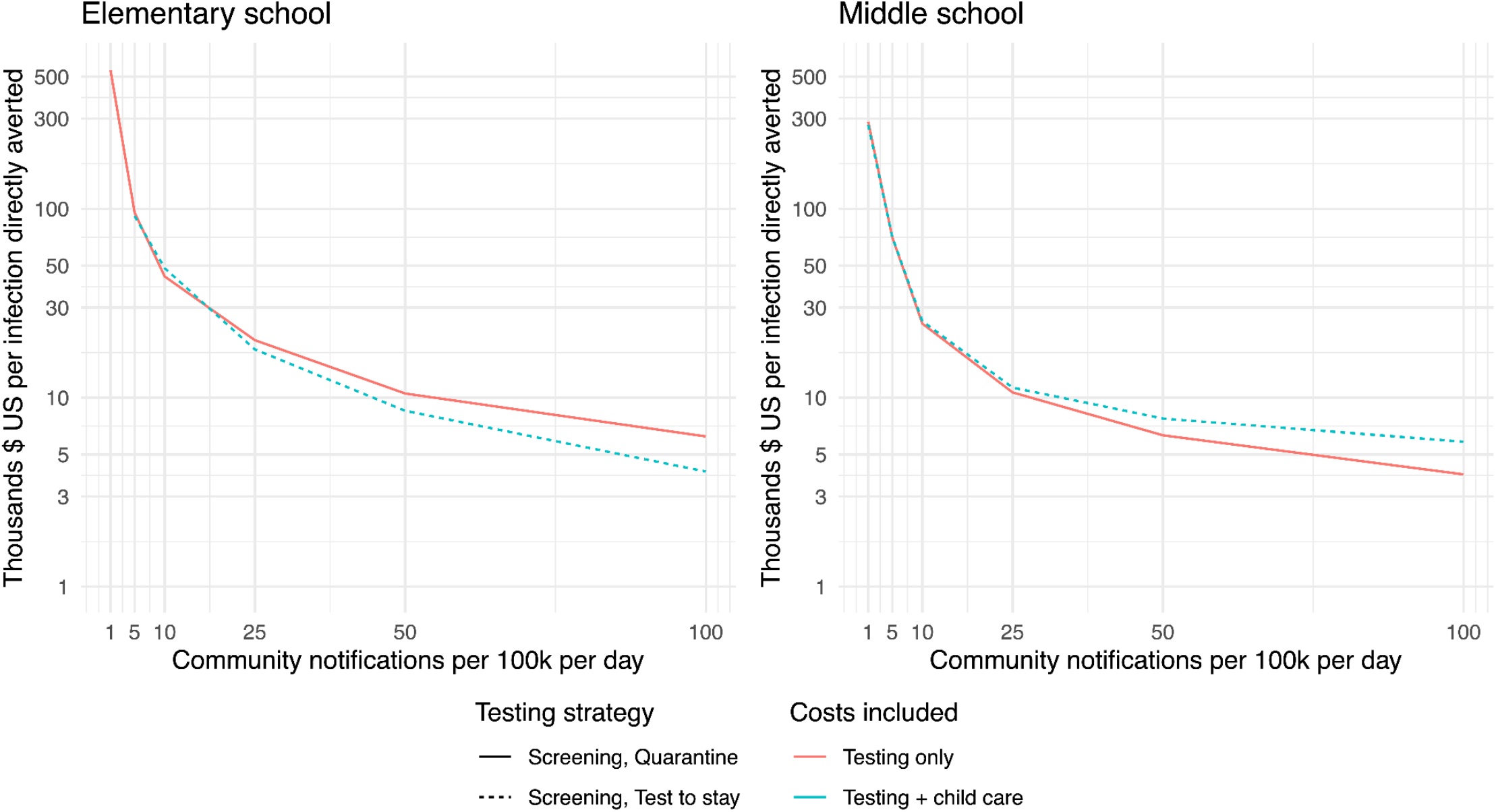
Cost per infection directly prevented among students/staff, compared to a 5-day in-person schedule with no in-school testing and high mitigation. Plots show the incremental cost, per infection directly averted among students and staff. For testing costs (orange), we show the strategy of weekly screening in which exposed contacts quarantine at home (solid line), which dominates the “test to stay” strategy. By “dominates”, we mean that if optimizing over test costs only, it is strictly higher value to quarantine contacts, rather than implement test-to-stay. Likewise, for combined costs of testing plus childcare (blue), we show the strategy of weekly screening with exposed contacts undergoing daily rapid tests to stay at school (dashed line), which dominates at-home quarantine. For alternative scenarios with rapid tests and/or lower in-school mitigation, see Figures S8 and S9.

### Sensitivity analysis

The infections averted by screening increased with higher true community COVID-19 incidence (including either higher community notification rates or lower community case detection rates) as well as with higher in-school attack rates (which could reflect weaker mitigation measures or greater variant transmissibility) (Figure S10). Correspondingly, higher attack rates reduced the cost of screening per infection averted by at least half in both settings (Figure S9). Screening later in the week or with a less sensitive test decreased impact slightly. Increasing testing frequency to twice per week increased the number of averted infections by <25% (Figure S10). For surveillance, reducing the fraction tested to 10% rather than 20% each week reduced impact but still allowed a response to large outbreaks; for example, it reduced the proportion of school-associated transmission prevented from 34% to 27% in a middle school at 10 community notifications/100k/day. Surveillance was more beneficial as the in-school attack rate increased (Figure S2).

## Discussion

Our work highlights that carefully designed COVID-19 testing can support safe school reopening and help maintain 5-day in-person education across a large range of community case rates. In particular, we underscore the importance of considering multiple dimensions of cost in school reopening plans. While school-based testing programs will increase expenditures, whether borne by school systems or supported by state or federal funding, these costs may be offset societally by reducing the burden of COVID-19-related childcare costs currently borne by parents and caregivers as well as costs associated with lost educational time.

Gains are particularly pronounced for expanded diagnostic testing, or “test to stay,” programs. We project that testing to stay results in only minor increases in transmission, even at the highest community case notification rates and with conservative assumptions about test sensitivity. Such estimates are consistent with a recent randomized controlled trial of “test to stay” programs in the United Kingdom, which were layered on top of twice-weekly screening programs (38). We further find that “test to stay” strategies have lower societal costs than quarantine-based strategies and could maintain student absences to less than 0.2% of school days. Given the current dominance of highly transmissible variants, an additional benefit of “test to stay” strategies is the option to adopt a broad definition of “close contact” without associated loss of school time.

We also provide information about the benefits and costs of two additional testing strategies: screening and surveillance. While previous analyses have documented that weekly screening can help control transmission, this analysis adds the finding that under conservative assumptions, 5-day in-person learning with screening is cost-saving from a societal perspective, compared to the hybrid or remote models often used in 2020-21 (39–41). Cost savings persist across levels of community transmission up to 100 cases/100k/day, even when improved case detection from the screening program increases the time that students spend in quarantine.

In 2020-21, screening was implemented in countries Germany, Austria, Norway, and the United Kingdom (18–22), as well as some US states (42,43), but its role in the coming year remains debated. We find that the value of screening varies substantially across different levels of community transmission, between elementary and middle schools, and by school attack rate. In turn, school attack rate is influenced by factors including mitigation measures (masking, ventilation, and distancing), vaccination uptake, and the properties of emerging variants of concern. As a result, screening capacity may be useful as an “insurance policy” to maintain in-personal instructional time if cases remain high during fall 2021, and would be most efficiently targeted toward older students, areas with low vaccination coverage, and settings where adherence to mitigation precautions is low or unknown.

In estimating impacts of testing on transmission, we did not include the downstream infections averted beyond students and staff, the medical costs associated with COVID-19 infection, or many other dimensions of cost (e.g., educational). Our estimates of cost per infection averted are therefore likely to be conservative, and when interpreting them, a school community’s willingness to pay per averted case should be affected by onward transmission risk. For example, setting the value per statistical life of $8 million (44), a common measure of willingness to pay, communities would be willing to invest $48,000 to avert a downstream infection in an unvaccinated person aged 50-64 and $720,000 per infection averted in those over 65 (45). Other important planning inputs might include local hospital capacity and any increased pediatric risks that may be associated with new variants. However, the availability of external federal funding may render the financial costs of testing less consequential for districts than logistical and practical considerations (19).

For districts concerned about in-school transmission, but without the capacity to perform regular screening, weekly surveillance of 10-20% of the school population may offer a middle ground. Surveillance (with the subsequent conversion to weekly screening when cases are identified) can reduce the risk of large outbreaks, is unlikely to falsely trigger burdensome interventions, and may allow schools to save money on testing when local incidence is low. However, surveillance of a small portion of the school population is likely to miss early outbreaks and requires regularly adapting school procedures. For these reasons, the benefit of surveillance strategies is largest when local testing is sparse (making it difficult to know how community case incidence maps to school incidence), when local incidence is rapidly changing, or when there is high uncertainty in the school attack rate. Beyond transmission impacts, both screening and surveillance also provide real-time information about case incidence in the school community, and may have value even at low incidence levels by providing reassurance to educators and parents concerned about in-person full-capacity attendance with the delta variant.

There are a number of limitations to this analysis. Guidance is still evolving in terms of who is a “close contact” in the context of new variants and which interventions are recommended for vaccinated individuals. Costs and benefits of testing strategies may change as recommendations evolve, but the ratio of testing compared to childcare costs should remain similar. In addition, our model does not address the operational aspects of specimen collection, laboratory transport, and reporting of results, which some schools have navigated successfully but may nevertheless pose barriers to adoption by others (19). Nevertheless, this work highlights that flexible and strategic testing can help ensure stable 5-day in-person education during the 2021-22 school year.

## Data Availability

Model code and replication files are publicly available as an R package on GitHub.

https://github.com/abilinski/BackToSchool2

## SUPPLEMENT

### Model Structure

We implemented a previously published SEIR model of COVID-19 transmission (1). Briefly, when individuals interacted with an agent (i.e. person) infected with SARS-CoV-2, transmission risk was proportional to duration and intensity of exposure. The model drew stochastic outcomes assuming an average incubation period of three days prior to the onset of infectiousness, two days of pre-symptomatic transmission if symptoms develop (2,3), total infectious time of five days (4–7), and overdispersion of infectivity in adolescents and adults (4,8) (Table 1). We assumed that adults with fully asymptomatic disease transmit COVID-19 at half the rate of those with any symptoms (9). Based on data from household contact tracing studies, we further specified that, in absence of vaccination, children under 10 were half as susceptible and half as infectious as symptomatic adolescents and adults (10–14). Beyond interactions with infectious agents within the simulation, students, staff, and their families had a probability of becoming infected through other community interactions equivalent to community per capita daily incidence assuming a 33% case detection rate. In vaccinated individuals, this risk was reduced by 80%; among unvaccinated adults, we upweighted community risk such that adults overall matched the community rate on average.

In scenarios without “test to stay”, symptom-driven COVID diagnostic testing still occurred outside of the school environment: individuals with COVID-19 who developed clinically-recognizable symptoms were assumed to self-isolate from out-of-household contacts (including staying home from school) and to obtain testing in the community. Results became available 24 hours after the first appearance of symptoms, at which point classrooms were notified and quarantined for 10 days. Symptom-driven community-based testing, and self-isolation of symptomatic individuals who had not been tested since symptom onset, were assumed to occur regardless of in-school screening practices.

For in-school testing, we assumed (a) specimens (e.g., anterior nasal swabs) were collected from each student and teacher, and (b) aliquots from up to eight specimens obtained in a single classroom were combined for pooled PCR testing, with negligible loss of sensitivity to detect active infection (15,16) When a pooled specimen yielded a positive result, all individual specimens that had been included in the pool were immediately tested separately using PCR to identify the positive individual(s).

#### Model Parameterization and Calibration

Model parameterization is discussed at length in the Supplement of (1). Briefly, we first identified household attack rates (including differential susceptibility and infectiousness of young children) (17– 19). We first adjusted these for the length of time spent in school and reduced infectiousness of asymptomatic individuals (9) to estimate attack rates with no or minimal mitigation. We then further adjusted them for a range of mitigation strategies. To partially validate our model, we compared our estimates of in-school attack rate and in-school Rt to those from empirical studies. We estimated in-school Rt with high mitigation and classroom quarantine and “bubbles” to be 0.2 for elementary schools and 0.64 in high schools, consistent with estimates from schools during 2020-2021 (e.g., (20)). Our estimates also reflect the wide range of attack rates across mitigation levels identified both in data directly from schools (21–23) and from household/population-level estimates (24,25), as well as the association between community incidence level and transmission risk (26).

We assumed that the delta variant is twice as transmissible than the wild type variant (27,28) and that this multiplicative increase is constant across levels of mitigation. The latter assumption is uncertain and requires further empirical evaluation in different contexts; for example, while it may be realistic with cloth masks, early anecdotal evidence from health care settings suggests that high filtration masks (e.g. N95, KN95) may protect nearly as well against the delta variant as they do against wild type. We also assumed that the delta variant has 80% vaccine efficacy, a decrease compared to the wild type (29,30). Some emerging estimates of vaccine efficacy are lower, potentially suggesting a conservative estimate of the value of screening.

For our base case, we assume high mitigation with the delta variant (Rt of approximately 0.4 in elementary schools and 1.2 in middle schools) to reflect the population of schools most likely to implement testing and likely ordering of interventions (e.g., testing will likely only be implemented in schools that have already implemented masking). We also present results with moderate mitigation, doubling the attack rate of the base case, to display the impact of testing in the context of reduced mitigation interventions in schools.

#### Surveillance Thresholds

Within a small school community, it is challenging to set an optimal threshold for triggering further investigation when conducting surveillance. We expect some COVID-19 cases to enter a school from the community *even if no transmission occurs within the school*, and ideally the threshold for triggering additional testing should take this into account. However, when testing a small fraction of the school (10-20%), the expected number of asymptomatic cases detected, assuming no in-school transmission, is generally close to 0. In the paper, we chose a 1-case trigger threshold for 3 reasons:

1. At low-to-moderate community notification rates (1-25 cases per 100,000/day), no surveillance scenario with a threshold above 1 could detect even large outbreaks of at least 10 in-school transmissions with any regularity: the maximum probability of detection (i.e., maximum sensitivity) was 35% for a 2-case threshold at 25 cases per 100,000/day. By contrast, a 1-case threshold had a detection probability of 30-75% across 1-25 cases per 100,000/day, while maintaining low rates of false positive triggers.
2. In our model, there was generally at least some in-school transmission at high levels of community incidence, making threshold selection less of a concern, since false positives would be rare under any threshold. (For the same reason, surveillance testing as a method of detecting outbreaks is less useful at these levels, although a benefit remains if community case detection is low, which makes schools less likely to be aware of local incidence risks.)
3. If, in practice, a school calibrated the expected number of cases and associated threshold to the *observed* community incidence rate, this would be a significant underestimate (and for most community incidence levels we evaluated would be near 0). (However, it is not straightforward to correct for case detection, as there is no public, consistently-collected data source in the United States for estimating case detection rate, and most school leaders with whom we spoke would not be comfortable making such an estimate.^1^)

Nevertheless, schools (or school districts more broadly) should adapt surveillance thresholds to meet their needs and level of caution. Our model is a stylized example over a single month for a single school. A longer-term strategy might include dynamic switching back to surveillance as well as stricter trigger thresholds when community incidence is high or when surveying large districts.

**Fig S1.**
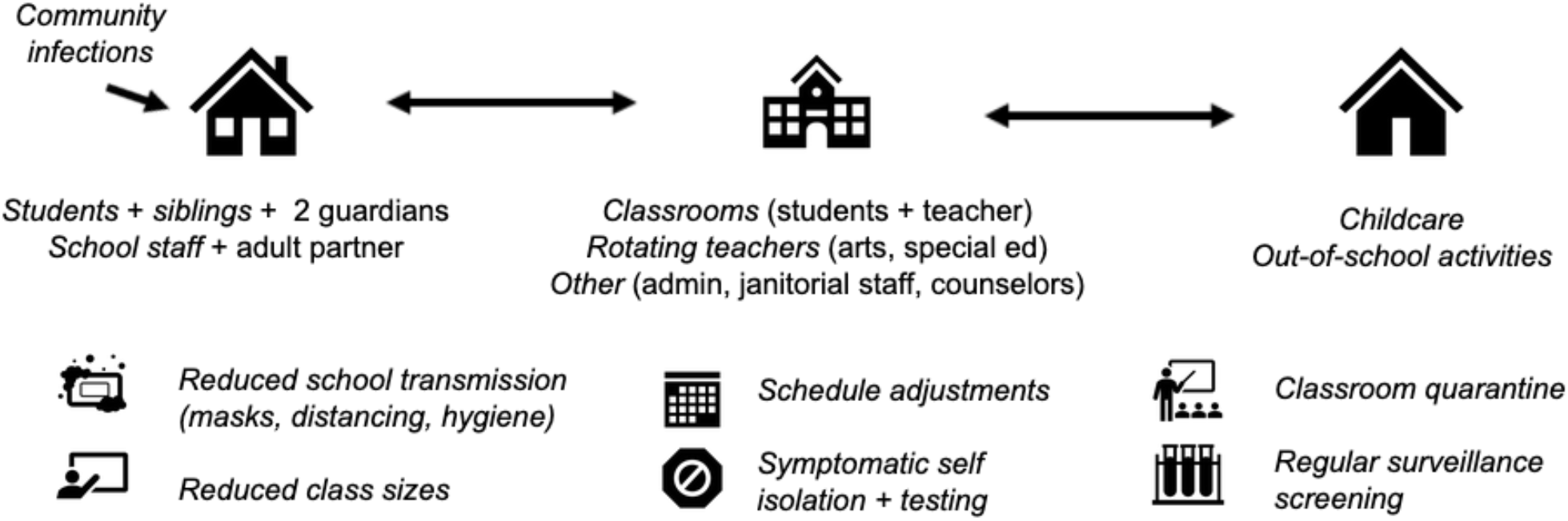
Model diagram.

**Fig S2.**
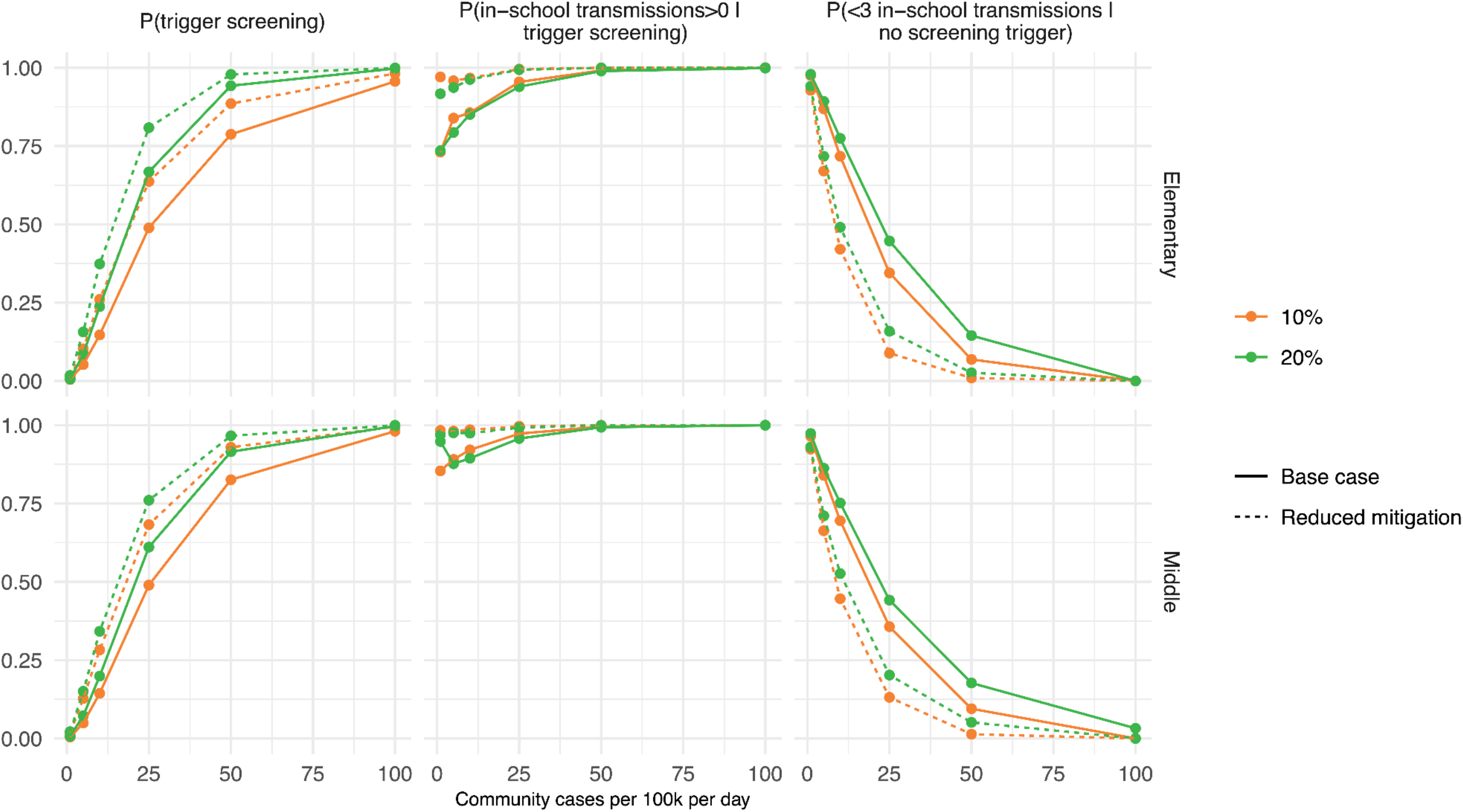
Surveillance characteristics. Color indicates the percentage of the school screened weekly (from unvaccinated individuals) under surveillance, while the line type indicates the transmission level. The left panels depict the probability of triggering screening. The middle panels depict the probability of in-school transmission, conditional on triggering screening (“true positives”). The right panels depict the probability of fewer than 3 in-school transmissions given no screening trigger (“true negatives”).

**Fig S3.**
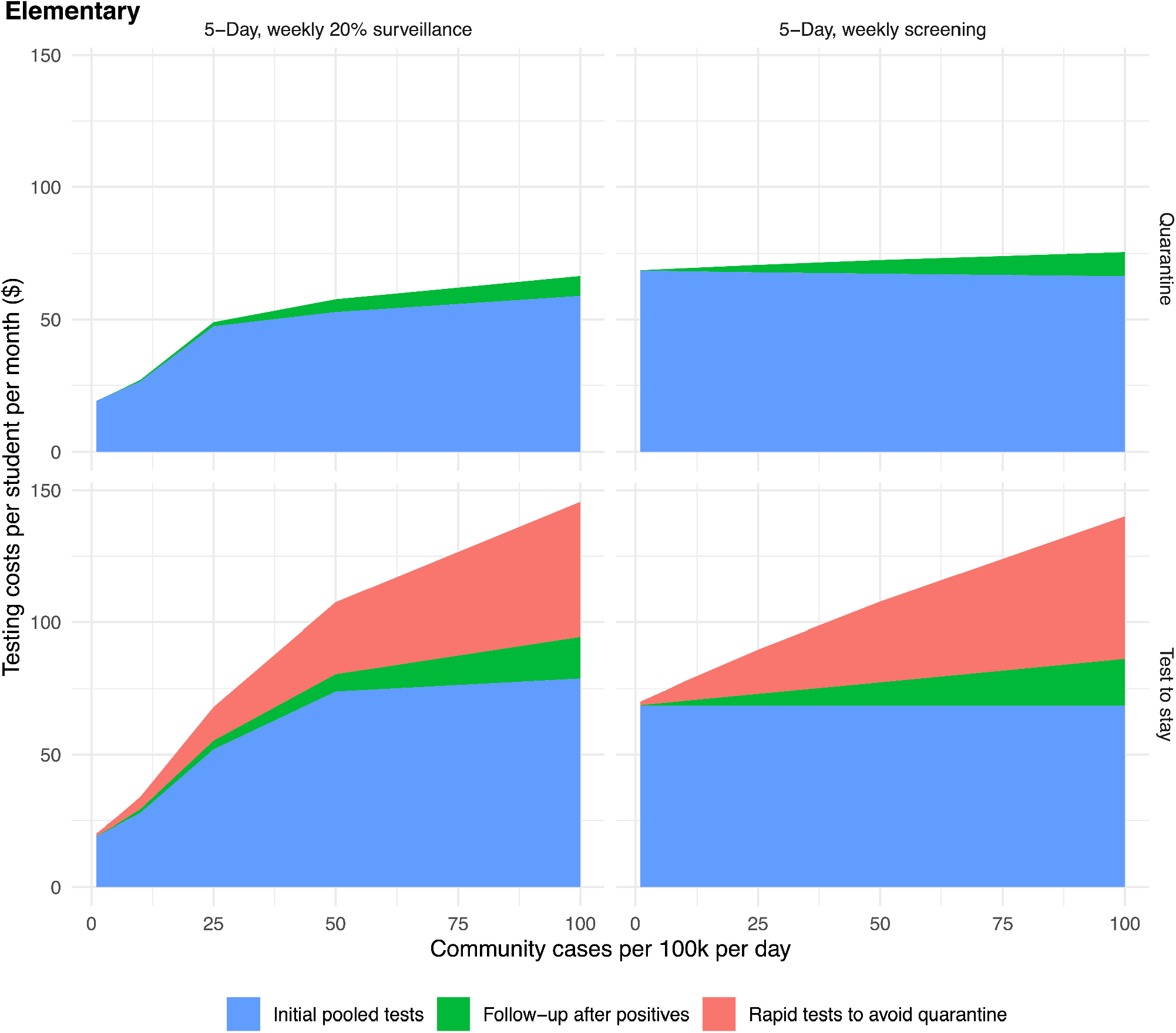
Testing costs, as dollars per student per month, in an elementary school. When exposed students quarantine at home, costs plateau at higher levels of incidence as classroom quarantines cause screening days to be missed; potential costs of community-based testing by exposed students or their contacts are not modeled. For a “test to stay” strategy that provides in-school rapid testing to symptomatic students and exposed contacts, testing costs increase as incidence rises.

**Fig S4.**
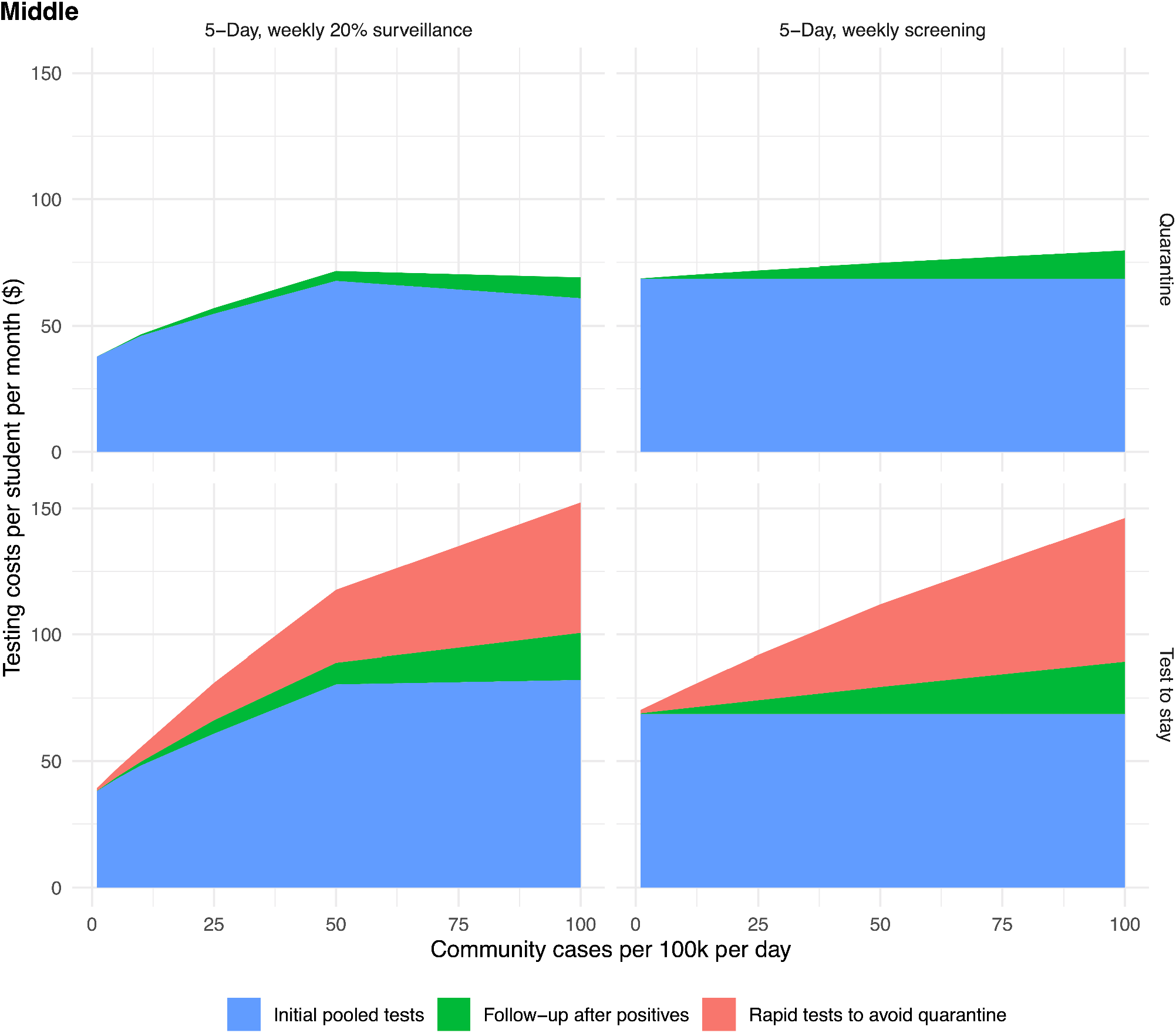
Testing costs, as dollars per student per month, in a middle school. When exposed students quarantine at home, costs plateau at higher levels of incidence as classroom quarantines cause screening days to be missed; potential costs of community-based testing by exposed students or their contacts are not modeled. For a “test to stay” strategy that provides in-school rapid testing to symptomatic students and exposed contacts, testing costs increase as incidence rises.

**Fig S5.**
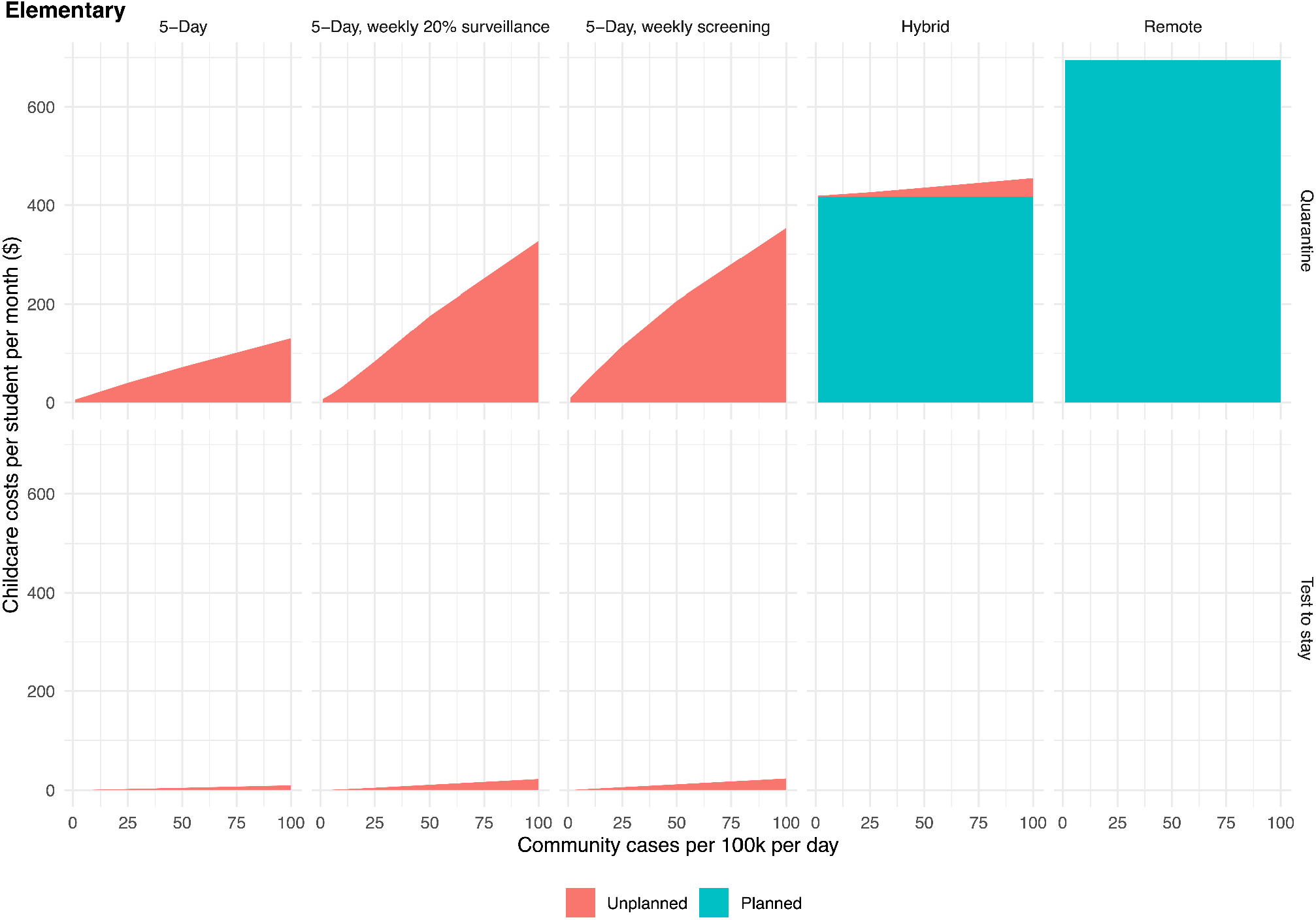
Childcare or parent productivity costs (elementary school). Planned costs reflect scheduled days of remote learning, and unplanned costs reflect days spent in isolation or quarantine. Rows reflect two different approaches to managing exposed contacts (quarantine for 10 days at home, top row; or staying at school with a week of daily rapid tests, bottom row). “Test to stay” is not modeled for Hybrid and Remote schedules.

**Fig S6.**
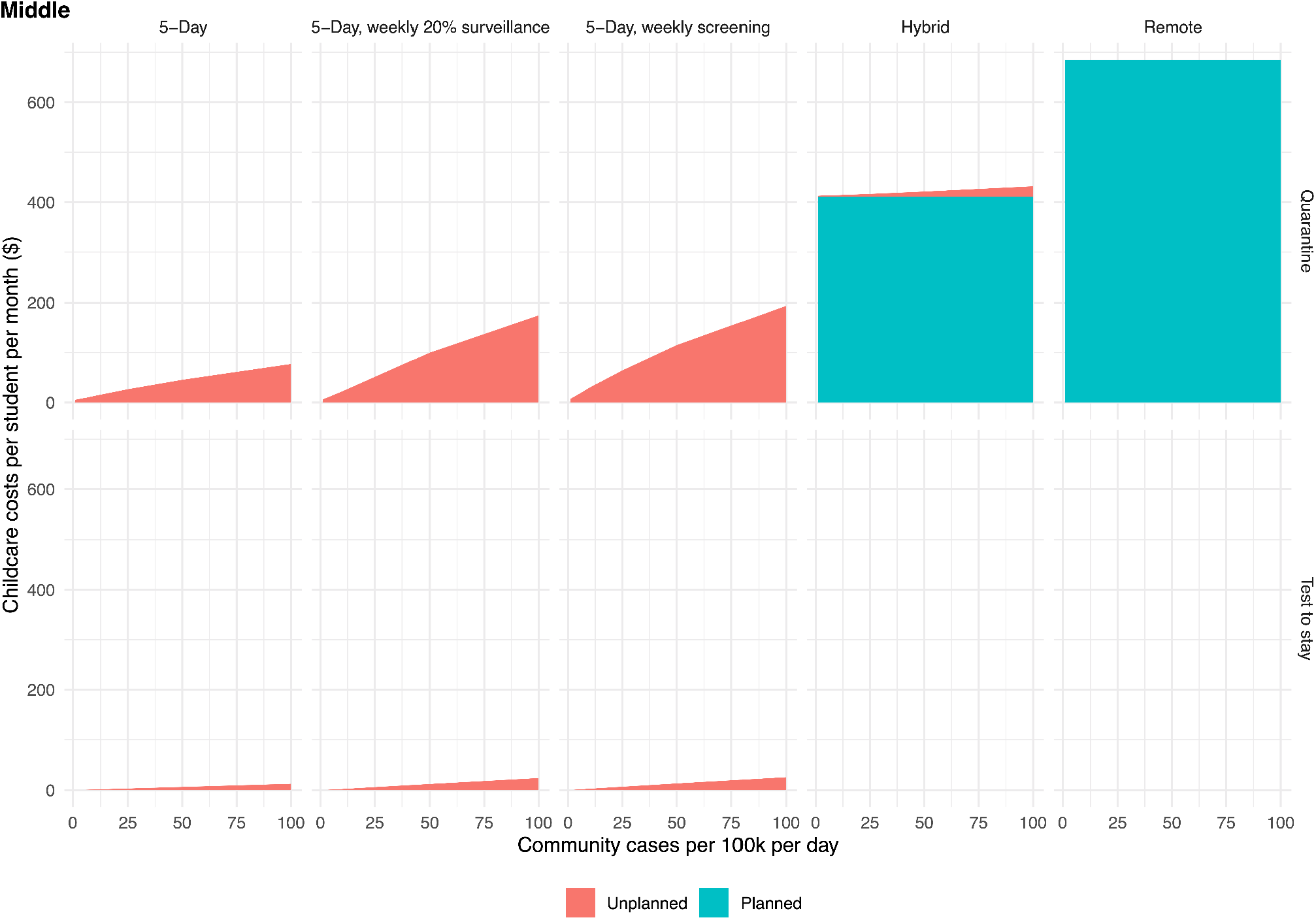
Childcare or parent productivity costs (middle school). We assume that for a combination of health and logistical reasons, full classrooms quarantine after exposure. If only unvaccinated students were asked to quarantine, then costs of 5-Day + Quarantine scenarios would be reduced.

**Fig S7.**
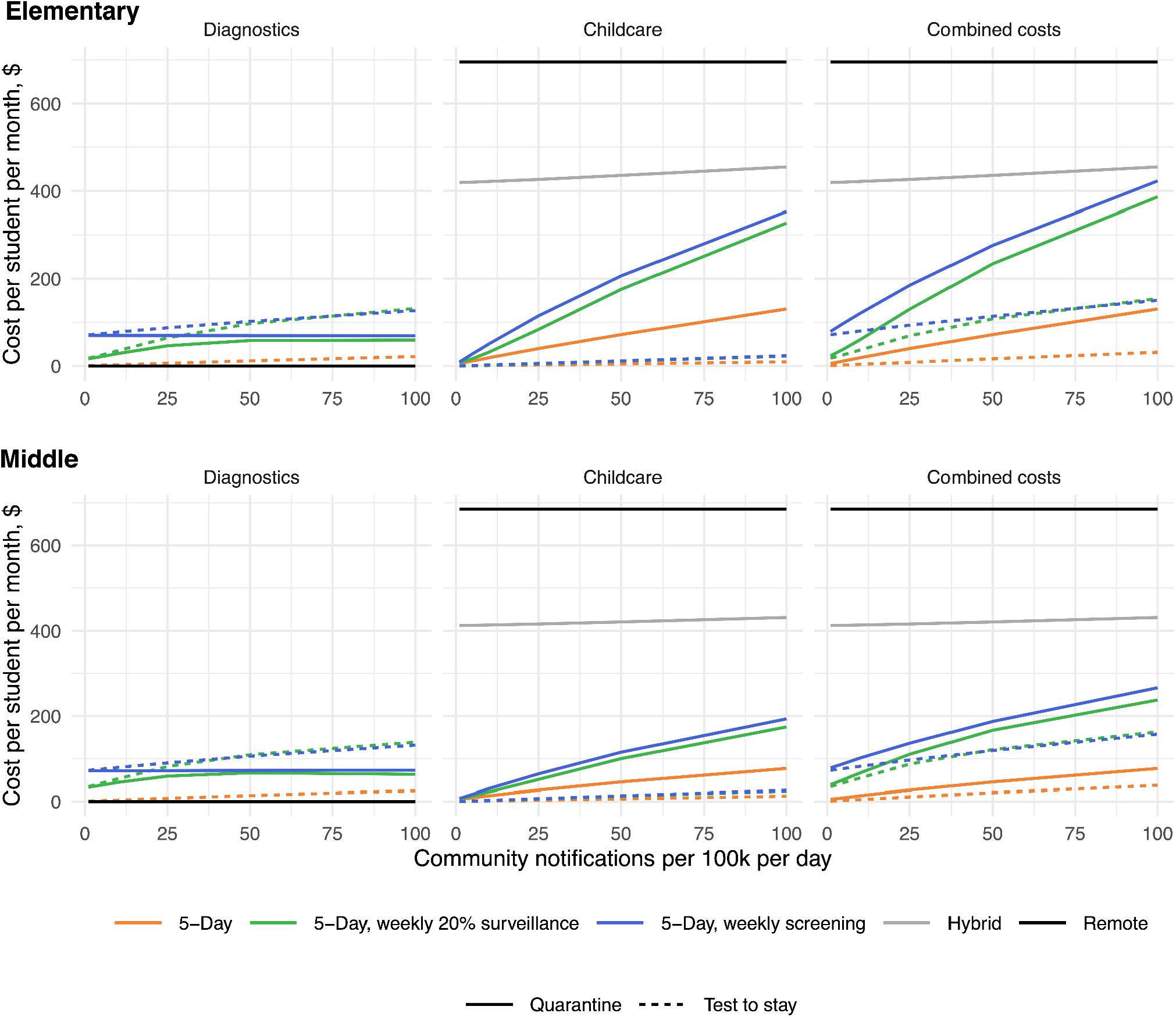
Costs associated with rapid antigen screening tests. (weekly tests at $12 per test + $8 per sample collection, PCR confirmation of positive results with same one-day turnaround, 0.5% false positive rapid tests, no change in sensitivity for acute infection) compared to the costs of schedule-based mitigation and of full-time in-person attendance without asymptomatic screening.

**Fig S8.**
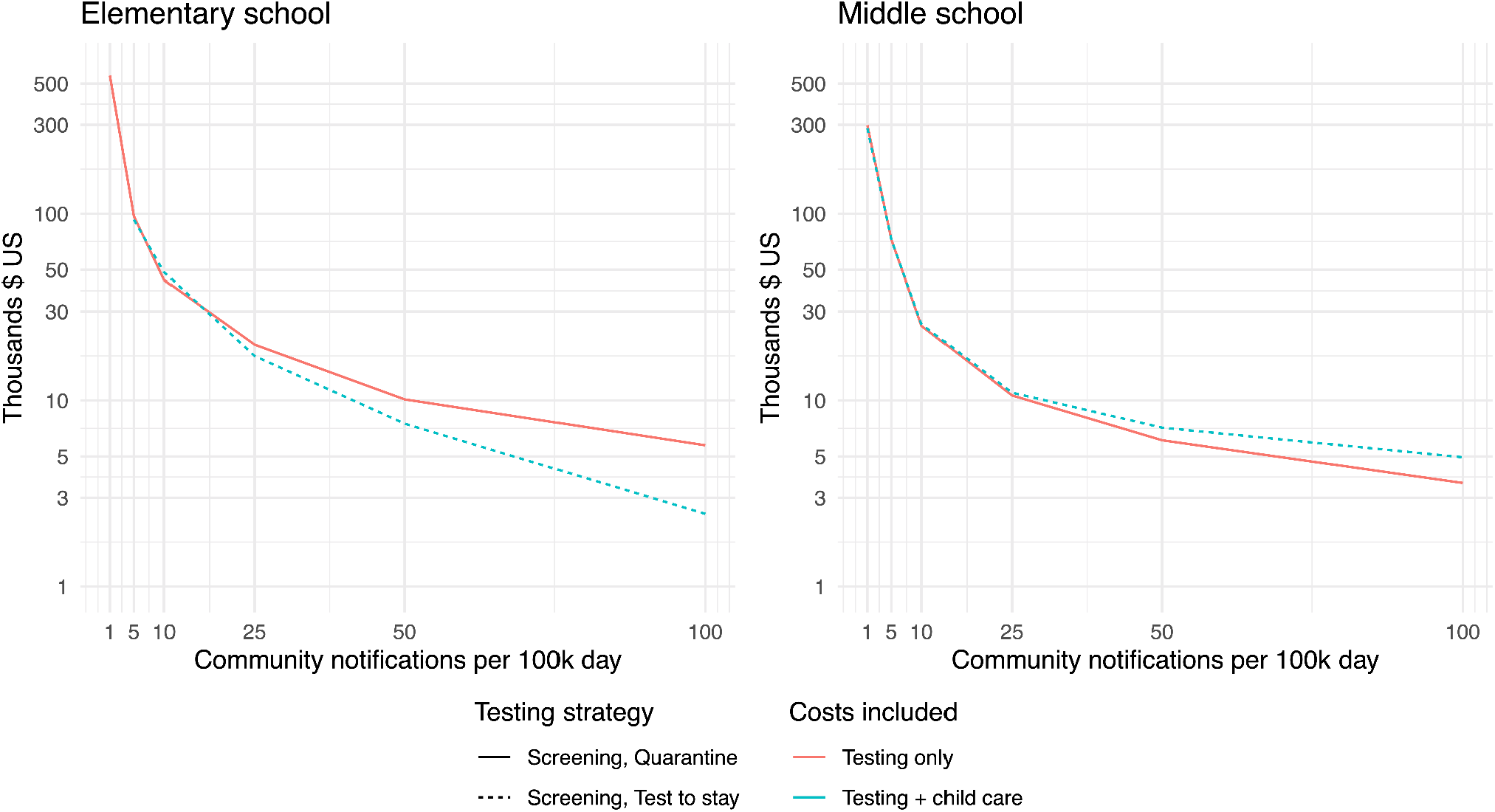
Cost-effectiveness of rapid screening (cost per infection directly averted among students and staff), comparing weekly screening to full-time attendance without screening, under the same rapid screening assumptions as in Figure S7. For testing costs (orange), we show the strategy of weekly screening in which exposed contacts quarantine at home (solid line), which dominates the “test to stay” strategy. By “dominates”, we mean that if optimizing over test costs only, it is strictly higher value to quarantine contacts, rather than implement test-to-stay. Likewise, for combined costs of testing plus childcare (blue), we show the strategy of weekly screening with exposed contacts undergoing daily rapid tests to stay at school (dashed line), which dominates at-home quarantine.

**Fig S9.**
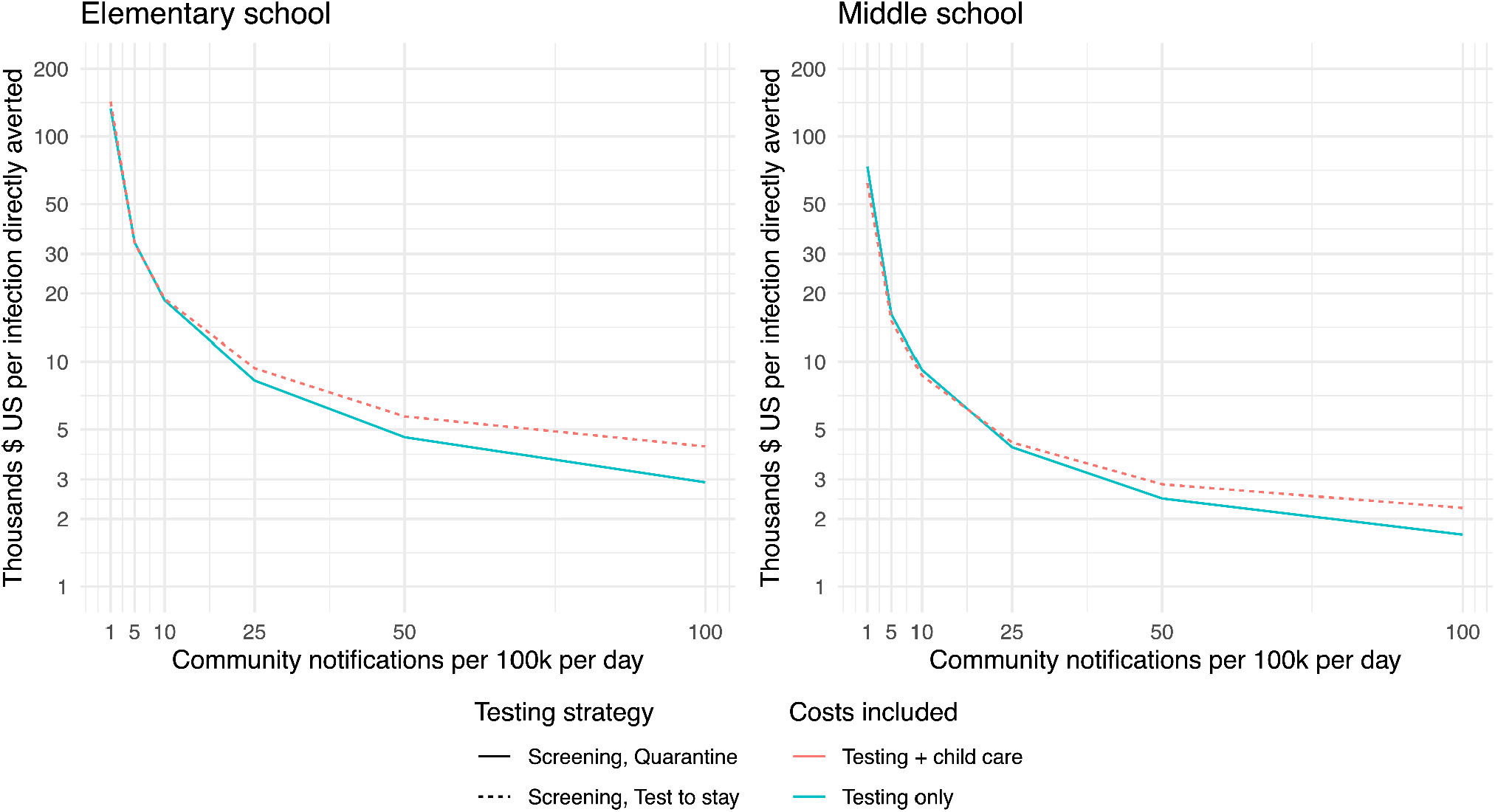
Cost-effectiveness of weekly screening (cost per infection directly averted among students and staff), comparing screening to full-time attendance without screening, assuming a two-fold increase in transmission rate over the base case (due to increased variant transmissibility or reduced in-school mitigation). For testing costs (orange), we show the strategy of weekly screening in which exposed contacts quarantine at home (solid line), which dominates the “test to stay” strategy. By “dominates”, we mean that if optimizing over test costs only, it is strictly higher value to quarantine contacts, rather than implement test-to-stay. Likewise, for combined costs of testing plus childcare (blue), we show the strategy of weekly screening with exposed contacts undergoing daily rapid tests to stay at school (dashed line), which dominates at-home quarantine.

**Fig S10.**
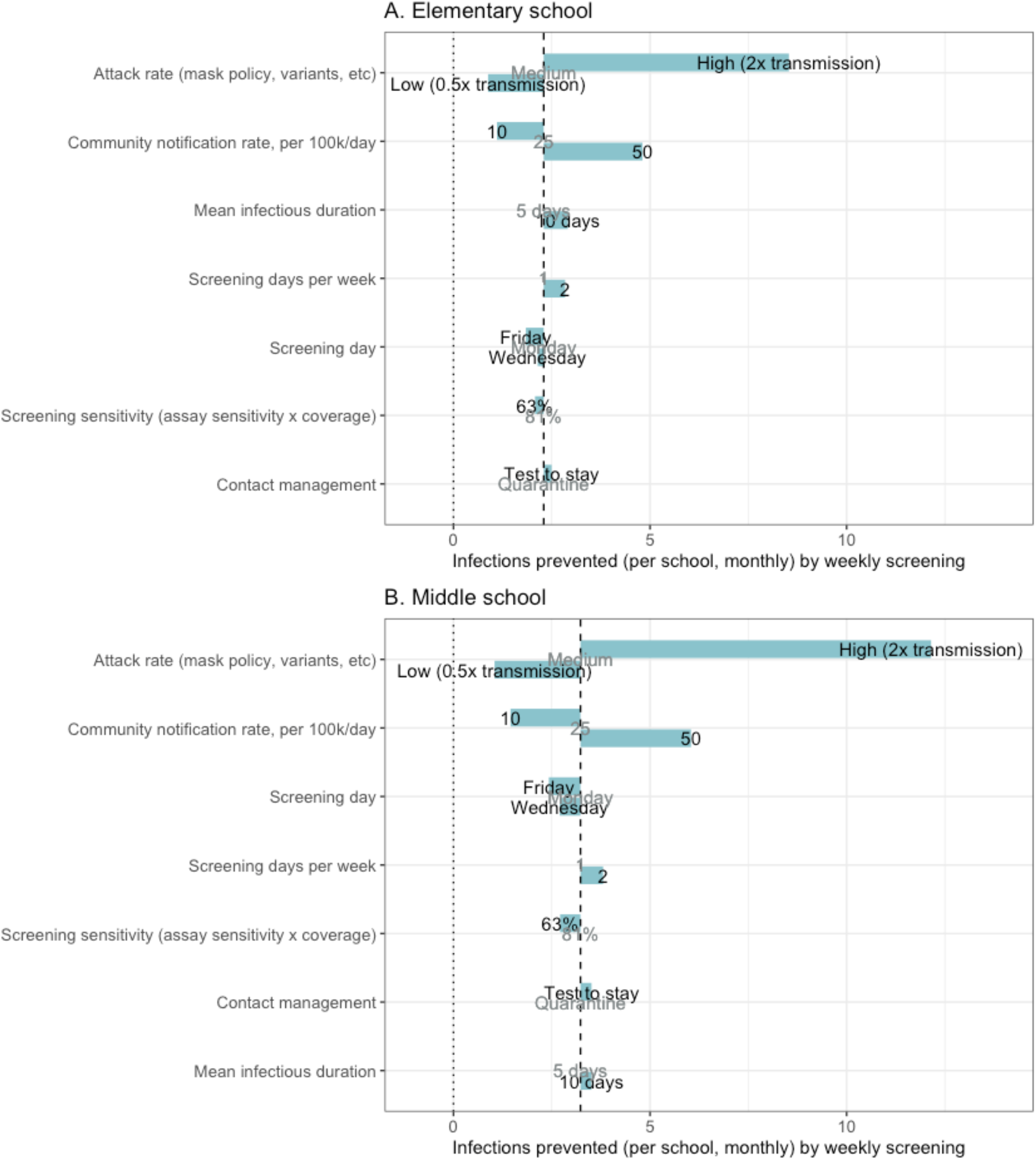
One-way sensitivity analyses, transmission effects of weekly screening. The outcome is the absolute difference in incidence (infections of students or teachers per school per month) between 5-day attendance with and without weekly screening, in an elementary (A) and middle school (B). The dotted horizontal line indicates the outcome when all parameters are at their base values (values indicated in gray text in the row where that parameter is varied).

**Table S1.** Comparison of transmission, case-detection, operational, and cost outcomes between different schedules and screening frequencies.

| | Infected (proportion of school per month) | Difference in proportion of school infected, per month vs full-time without screening | Proportion of incremental infections prevented (of difference between 5-day no screening and Remote) | Proportion of cases detected | In-person attendance (proportion of school days) | Testing costs (\$ per student per month) | Testing + child care costs (\$ per student per month) |
| --- | --- | --- | --- | --- | --- | --- | --- |
| Elementary school, community notification rate 10/100k/day |  |  |  |  |  |  |  |
| **5-Day, no screening, quarantine** | 0.01 | 0 | 0 | 0.23 | 0.989 | 0 | 19.03 |
| **5-Day, no screening, test-to-stay** | 0.01 | 0.0002 | -0.06 | 0.23 | 0.999 | 3.47 | 4.36 |
| **5-Day, weekly 10% surveillance, quarantine** | 0.009 | -0.0005 | 0.19 | 0.31 | 0.985 | 18.93 | 44.94 |
| **5-Day, weekly 10% surveillance, test-to-stay** | 0.01 | -0.0002 | 0.06 | 0.35 | 0.999 | 24.25 | 25.56 |
| **5-Day, weekly 20% surveillance, quarantine** | 0.009 | -0.0007 | 0.25 | 0.39 | 0.981 | 27.09 | 58.97 |
| **5-Day, weekly 20% surveillance, test-to-stay** | 0.009 | -0.0005 | 0.17 | 0.44 | 0.999 | 33.98 | 35.59 |
| **5-Day, 1x/week screening, quarantine** | 0.008 | -0.0016 | 0.57 | 0.66 | 0.97 | 69.42 | 120.26 |
| **5-Day, 1x/week screening, test-to-stay** | 0.009 | -0.0013 | 0.46 | 0.73 | 0.999 | 77.81 | 80.25 |
| **5-Day, 2x/week screening, quarantine** | 0.008 | -0.0019 | 0.69 | 0.77 | 0.968 | 124.19 | 178.24 |
| **5-Day, 2x/week weekly screening, test-to-stay** | 0.008 | -0.0017 | 0.61 | 0.88 | 0.998 | 134.03 | 136.86 |
| **Hybrid** | 0.007 | -0.0025 | 0.92 | 0.23 | 0.397 | 0 | 421.76 |
| **Remote** | 0.007 | -0.0028 | 1 | 0 | 0 | 0 | 695.32 |
| **Elementary school, community notification rate 50/100k/day** |  |  |  |  |  |  |  |
| **5-Day, no screening, quarantine** | 0.046 | 0 | 0 | 0.22 | 0.957 | 0 | 71.99 |
| **5-Day, no screening, test-to-stay** | 0.047 | 0.0013 | -0.1 | 0.25 | 0.997 | 14.73 | 19.41 |
| **5-Day, weekly 10% surveillance, quarantine** | 0.042 | -0.0034 | 0.28 | 0.45 | 0.912 | 46.65 | 194.49 |
| **5-Day, weekly 10% surveillance, test-to-stay** | 0.043 | -0.0026 | 0.21 | 0.55 | 0.994 | 80.29 | 89.69 |
| **5-Day, weekly 20% surveillance, quarantine** | 0.041 | -0.0054 | 0.44 | 0.54 | 0.896 | 57.65 | 233.33 |
| **5-Day, weekly 20% surveillance, test-to-stay** | 0.042 | -0.0038 | 0.32 | 0.66 | 0.994 | 107.6 | 118.43 |
| **5-Day, 1x/week screening, quarantine** | 0.039 | -0.0069 | 0.57 | 0.63 | 0.877 | 72.52 | 278.98 |
| **5-Day, 1x/week screening, test-to-stay** | 0.04 | -0.0056 | 0.46 | 0.74 | 0.993 | 107.84 | 119.56 |
| **5-Day, 2x/week screening, quarantine** | 0.038 | -0.0084 | 0.69 | 0.73 | 0.87 | 126.85 | 345.93 |
| **5-Day, 2x/week weekly screening, test-to-stay** | 0.039 | -0.0064 | 0.53 | 0.89 | 0.992 | 169.93 | 183.79 |
| **Hybrid** | 0.036 | -0.0099 | 0.81 | 0.22 | 0.389 | 0 | 435.62 |
| **Remote** | 0.034 | -0.0121 | 1 | 0 | 0 | 0 | 695.32 |
| **Middle school, community notification rate 10/100k/day** |  |  |  |  |  |  |  |
| **5-Day, no screening, quarantine** | 0.012 | 0 | 0 | 0.23 | 0.992 | 0 | 13.48 |
| **5-Day, no screening, test-to-stay** | 0.013 | 0.0006 | -0.12 | 0.26 | 0.999 | 46.62 | 68.93 |
| **5-Day, weekly 10% surveillance, quarantine** | 0.011 | -0.0013 | 0.27 | 0.34 | 0.989 | 4.47 | 5.76 |
| **5-Day, weekly 10% surveillance, test-to-stay** | 0.011 | -0.0008 | 0.16 | 0.42 | 0.999 | 55.46 | 57.6 |
| **5-Day, weekly 20% surveillance, quarantine** | 0.011 | -0.0017 | 0.34 | 0.43 | 0.987 | 23.84 | 42.07 |
| **5-Day, weekly 20% surveillance, test-to-stay** | 0.011 | -0.0015 | 0.3 | 0.52 | 0.999 | 31.24 | 33.09 |
| **5-Day, 1x/week screening, quarantine** | 0.009 | -0.0028 | 0.57 | 0.64 | 0.982 | 70.06 | 100.16 |
| **5-Day, 1x/week screening, test-to-stay** | 0.01 | -0.0027 | 0.54 | 0.74 | 0.998 | 78.57 | 81.31 |
| **5-Day, 2x/week screening, quarantine** | 0.009 | -0.0035 | 0.7 | 0.77 | 0.982 | 125.35 | 155.67 |
| **5-Day, 2x/week weekly screening, test-to-stay** | 0.009 | -0.0031 | 0.62 | 0.9 | 0.998 | 135.35 | 138.51 |
| **Hybrid** | 0.008 | -0.0044 | 0.89 | 0.22 | 0.398 | 0 | 413.93 |
| **Remote** | 0.007 | -0.005 | 1 | 0 | 0 | 0 | 684.79 |
| **Middle school, community notification rate 50/100k/day** |  |  |  |  |  |  |  |
| **5-Day, no screening, quarantine** | 0.055 | 0 | 0 | 0.22 | 0.973 | 0 | 45.46 |
| **5-Day, no screening, test-to-stay** | 0.058 | 0.0023 | -0.11 | 0.28 | 0.996 | 71.78 | 171.71 |
| **5-Day, weekly 10% surveillance, quarantine** | 0.049 | -0.0062 | 0.31 | 0.44 | 0.947 | 18.18 | 24.48 |
| **5-Day, weekly 10% surveillance, test-to-stay** | 0.051 | -0.0049 | 0.24 | 0.59 | 0.993 | 117.73 | 129.93 |
| **5-Day, weekly 20% surveillance, quarantine** | 0.047 | -0.0089 | 0.44 | 0.51 | 0.94 | 52.13 | 139.47 |
| **5-Day, weekly 20% surveillance, test-to-stay** | 0.048 | -0.0073 | 0.36 | 0.66 | 0.993 | 92.72 | 104.17 |
| **5-Day, 1x/week screening, quarantine** | 0.044 | -0.0118 | 0.58 | 0.63 | 0.931 | 74.8 | 189.83 |
| **5-Day, 1x/week screening, test-to-stay** | 0.045 | -0.0103 | 0.5 | 0.76 | 0.992 | 111.96 | 125.28 |
| **5-Day, 2x/week screening, quarantine** | 0.041 | -0.0148 | 0.73 | 0.75 | 0.929 | 131.25 | 248.46 |
| **5-Day, 2x/week weekly screening, test-to-stay** | 0.043 | -0.0123 | 0.6 | 0.9 | 0.991 | 173.31 | 188.37 |
| **Hybrid** | 0.037 | -0.0183 | 0.9 | 0.23 | 0.394 | 0 | 421.16 |
| **Remote** | 0.035 | -0.0204 | 1 | 0 | 0 | 0 | 684.79 |

## Footnotes

1 Comparing percent positivity from in-school testing to population testing is inappropriate, as community testing encompasses primarily exposed symptomatic individuals with a much higher probability of infection than randomly selected individuals.

## References

1. Burbio School and Community Events Data Platform [Internet]. [cited 2021 Feb 22]. Available from: https://info.burbio.com/school-tracker-update-feb-22/

2. 2020-2021 Boston Public Schools Dashboard [Internet]. Google Data Studio. [cited 2021 Apr 21]. Available from: http://datastudio.google.com/reporting/77e82eef-349a-4f00-bd50-0824cd0df332/page/1vChB?feature=opengraph

3. Shapiro E. Over 50,000 N.Y.C. public school students will return to classrooms, including in middle and high school. The New York Times [Internet]. Y2021 Apr 12 [cited 2021 Apr 21]; Available from: https://www.nytimes.com/2021/04/12/nyregion/nyc-public-schools-students.html

4. Polikoff AR Anna Saavedra, Dan Silver, and Morgan. Surveys show things are better for students than they were in the spring—or do they? [Internet]. Brookings. 2020 [cited 2020 Nov 20]. Available from: https://www.brookings.edu/blog/brown-center-chalkboard/2020/11/18/surveys-show-things-are-better-for-students-than-they-were-in-the-spring-or-do-they/

5. Gaudiano N. Missing: Millions of students [Internet]. POLITICO. [cited 2020 Nov 25]. Available from: https://politi.co/3dVrNxg

6. Leeb RT. Mental Health–Related Emergency Department Visits Among Children Aged 18 Years During the COVID-19 Pandemic — United States, January 1–October 17, 2020. MMWR Morb Mortal Wkly Rep [Internet]. 2020 [cited 2020 Nov 30];69. Available from: https://www.cdc.gov/mmwr/volumes/69/wr/mm6945a3.htm

7. Hess A. Widespread school closures mean 30 million kids might go without meals [Internet]. CNBC. 2020 [cited 2020 Aug 1]. Available from: https://www.cnbc.com/2020/03/14/widespread-school-closures-mean-30-million-kids-might-go-without-meals.html

8. Baron EJ, Goldstein EG, Wallace CT. Suffering in Silence: How COVID-19 School Closures Inhibit the Reporting of Child Maltreatment [Internet]. Rochester, NY: Social Science Research Network; 2020 Jul [cited 2020 Aug 1]. Report No.: ID 3601399. Available from: https://papers.ssrn.com/abstract=3664803

9. Bueno C. Bricks and Mortar vs. Computers and Modems: The Impacts of Enrollment in K-12 Virtual Schools [Internet]. Rochester, NY: Social Science Research Network; 2020 Jul [cited 2020 Aug 1]. Report No.: ID 3642969. Available from: https://papers.ssrn.com/abstract=3642969

10. Soland J, Kuhfeld M, Tarasawa B, Johnson A, Ruzek E, Liu J. The impact of COVID-19 on student achievement and what it may mean for educators [Internet]. Brookings. 2020 [cited 2020 Aug 1]. Available from: https://www.brookings.edu/blog/brown-center-chalkboard/2020/05/27/the-impact-of-covid-19-on-student-achievement-and-what-it-may-mean-for-educators/

11. Goldhaber D, Imberman S, Strunk KO, Hopkins B, Brown N, Harbatkin E, et al. To What Extent Does In-Person Schooling Contribute to the Spread of COVID-19? Evidence from Michigan and Washington [Internet]. 2020 Dec [cited 2021 Jan 24]. Available from: https://epicedpolicy.org/does-in-person-schooling-contribute-to-the-spread-of-covid-19/

12. Harris D, Ziedan E, Hassig S. The Effects of School Reopenings on COVID-19 Hospitalizations [Internet]. [cited 2021 Jan 24]. Available from: https://www.reachcentered.org/publications/the-effects-of-school-reopenings-on-covid-19-hospitalizations

13. Volz E, Mishra S, Chand M, Barrett JC, Johnson R, Geidelberg L, et al. Transmission of SARS-CoV-2 Lineage B.1.1.7 in England: Insights from linking epidemiological and genetic data. medRxiv. 2021 Jan 4;2020.12.30.20249034.

14. Panovska-Griffiths J, Kerr C, Stuart RM, Mistry D, Klein D, Viner RM, et al. Determining the optimal strategy for reopening schools, work and society in the UK: balancing earlier opening and the impact of test and trace strategies with the risk of occurrence of a secondary COVID-19 pandemic wave. medRxiv. 2020 Jun 1;2020.06.01.20100461.

15. Schechter-Perkins EM, van den Berg P, Branch-Elliman W. The Science Behind Safe School Re-Opening: Leveraging the Pillars of Infection Control to Support Safe Elementary and Secondary Education During the COVID-19 Pandemic. Open Forum Infectious Diseases [Internet]. 2021 Mar 17 [cited 2021 Apr 29];(ofab134). Available from: https://doi.org/10.1093/ofid/ofab134

16. CDC. Guidance for COVID-19 Prevention in K-12 Schools and ECE Programs [Internet]. Centers for Disease Control and Prevention. 2021.[cited 2021 Jul 19]. Available from: https://www.cdc.gov/coronavirus/2019-ncov/community/schools-childcare/k-12-guidance.html

17. White House Announces $10 Billion For COVID-19 Testing In Schools [Internet]. NPR.org. [cited 2021 May 2]. Available from: https://www.npr.org/sections/coronavirus-live-updates/2021/03/17/978262865/white-house-announces-10-billion-for-covid-19-testing-in-schools

18. Doron S, Ingalls RR, Beauchamp A, Boehm J, Boucher HW, Chow LH, et al. Weekly SARS-CoV-2 screening of asymptomatic students and staff to guide and evaluate strategies for safer in-person learning. medRxiv. 2021 Mar 22;2021.03.20.21253976.

19. Ciaranello A, Goehringer C, Nelson SB, Ruark LJ, Pollock NR. Lessons learned from implementation of SARS-CoV-2 screening in K-12 public schools in Massachusetts. Open Forum Infectious Diseases [Internet]. 2021 Jun 4 [cited 2021 Aug 3];(ofab287). Available from: https://doi.org/10.1093/ofid/ofab287

20. CDC. Communities, Schools, Workplaces, & Events [Internet]. Centers for Disease Control and Prevention. 2020.[cited 2020 Nov 25]. Available from: https://www.cdc.gov/coronavirus/2019-ncov/community/schools-childcare/indicators.html

21. COVID-19 Vaccination | CDC [Internet]. 2021.[cited 2021 Apr 20]. Available from: https://www.cdc.gov/vaccines/covid-19/index.html

22. Bilinski A, Salomon JA, Giardina J, Ciaranello A, Fitzpatrick MC. Passing the Test: A Model-Based Analysis of Safe School-Reopening Strategies. Ann Intern Med [Internet]. 2021 Jun 8 [cited 2021 Aug 9]; Available from: https://www.acpjournals.org/doi/10.7326/M21-0600

23. Wheaton WD. U.S. Synthetic Population 2010 Version 1.0 Quick Start Guide, RTI International. 2014 May.

24. Fontanet A, Grant R, Tondeur L, Madec Y, Grzelak L, Cailleau I, et al. SARS-CoV-2 infection in primary schools in northern France: A retrospective cohort study in an area of high transmission. medRxiv. 2020 Jun 29;2020.06.25.20140178.

25. Stein-Zamir C, Abramson N, Shoob H, Libal E, Bitan M, Cardash T, et al. A large COVID-19 outbreak in a high school 10 days after schools’ reopening, Israel, May 2020. Eurosurveillance. 2020 Jul 23;25(29):2001352.

26. Dattner I, Goldberg Y, Katriel G, Yaari R, Gal N, Miron Y, et al. The role of children in the spread of COVID-19: Using household data from Bnei Brak, Israel, to estimate the relative susceptibility and infectivity of children [Internet]. Infectious Diseases (except HIV/AIDS); 2020 Jun [cited 2020 Jun 19]. Available from: http://medrxiv.org/lookup/doi/10.1101/2020.06.03.20121145

27. Park YJ, Choe YJ, Park O, Park SY, Kim Y-M, Kim J, et al. Early Release - Contact Tracing during Coronavirus Disease Outbreak, South Korea, 2020 - Volume 26, Number 10—October 2020 - Emerging Infectious Diseases journal - CDC. [cited 2020 Aug 1]; Available from: https://www.nc.cdc.gov/eid/article/26/10/20-1315_article

28. Fontanet A, Tondeur L, Madec Y, Grant R, Besombes C, Jolly N, et al. Cluster of COVID-19 in northern France: A retrospective closed cohort study. medRxiv. 2020 Apr 23;2020.04.18.20071134.

29. CDC. Delta Variant: What We Know About the Science [Internet]. Centers for Disease Control and Prevention. 2021 [cited 2021 Aug 9]. Available from: https://www.cdc.gov/coronavirus/2019-ncov/variants/delta-variant.html

30. Campbell F, Archer B, Laurenson-Schafer H, Jinnai Y, Konings F, Batra N, et al. Increased transmissibility and global spread of SARS-CoV-2 variants of concern as at June 2021. Eurosurveillance. 2021 Jun 17;26(24):2100509.

31. Sheikh A, McMenamin J, Taylor B, Robertson C, Public Health Scotland and the EAVE II Collaborators. SARS-CoV-2 Delta VOC in Scotland: demographics, risk of hospital admission, and vaccine effectiveness. Lancet. 2021 Jun 26;397(10293):2461–2.

32. Lopez Bernal J, Andrews N, Gower C, Gallagher E, Simmons R, Thelwall S, et al. Effectiveness of Covid-19 Vaccines against the B.1.617.2 (Delta) Variant. New England Journal of Medicine. 2021 Jul 21;0(0):null.

33. Duggan ME.

34. Lanier WA. COVID-19 Testing to Sustain In-Person Instruction and Extracurricular Activities in High Schools — Utah, November 2020–March 2021. MMWR Morb Mortal Wkly Rep [Internet]. 2021 [cited 2021 Jun 22];70. Available from: https://www.cdc.gov/mmwr/volumes/70/wr/mm7021e2.htm

35. Prince-Guerra JL. Evaluation of Abbott BinaxNOW Rapid Antigen Test for SARS-CoV-2 Infection at Two Community-Based Testing Sites — Pima County, Arizona, November 3–17, 2020. MMWR Morb Mortal Wkly Rep [Internet]. 2021 [cited 2021 Apr 25];70. Available from: https://www.cdc.gov/mmwr/volumes/70/wr/mm7003e3.htm

36. Mina MJ, Parker R, Larremore DB. Rethinking Covid-19 Test Sensitivity — A Strategy for Containment. New England Journal of Medicine. 2020 Nov 26;383(22):e120.

37. Childcare Workers [Internet]. 2020 [cited 2021 Apr 20]. Available from: https://www.bls.gov/oes/current/oes399011.htm

38. Young BC, Eyre DW, Kendrick S, White C, Smith S, Beveridge G, et al. A cluster randomised trial of the impact of a policy of daily testing for contacts of COVID-19 cases on attendance and COVID-19 transmission in English secondary schools and colleges [Internet]. Infectious Diseases (except HIV/AIDS); 2021 Jul [cited 2021 Aug 3]. Available from: http://medrxiv.org/lookup/doi/10.1101/2021.07.23.21260992

39. Bilinski A, Salomon JA, Giardina J, Ciaranello A, Fitzpatrick MC. Passing the Test: A Model-based analysis of safe school-reopening strategies. medRxiv. 2021 Jan 29;2021.01.27.21250388.

40. McGee RS, Homburger JR, Williams HE, Bergstrom CT, Zhou AY. Model-driven mitigation measures for reopening schools during the COVID-19 pandemic. medRxiv. 2021 Feb 6;2021.01.22.21250282.

41. Gill BP, Goyal R, Hotchkiss J. Operating Schools in a Pandemic: Predicted Effects of Opening, Quarantining, and Closing Strategies. :63.

42. Coronavirus/COVID-19: Pooled Testing in K-12 Schools [Internet]. [cited 2021 Feb 22]. Available from: https://www.doe.mass.edu/covid19/pooled-testing/

43. COVID Testing Results [Internet]. web. [cited 2021 Jan 24]. Available from: https://www.schools.nyc.gov/school-year-20-21/return-to-school-2020/health-and-safety/covid-19-testing/covid-testing-results

44. Robinson LA, Sullivan R, Shogren JF. Do the Benefits of COVID-19 Policies Exceed the Costs? Exploring Uncertainties in the Age–VSL Relationship. Risk Anal. 2020 Jul 16;10.1111/risa.13561.

45. CDC. COVID-19 Pandemic Planning Scenarios [Internet]. Centers for Disease Control and Prevention. 2020 [cited 2021 Aug 10]. Available from: https://www.cdc.gov/coronavirus/2019-ncov/hcp/planning-scenarios.html

46. Alene M, Yismaw L, Assemie MA, Ketema DB, Gietaneh W, Birhan TY. Serial interval and incubation period of COVID-19: a systematic review and meta-analysis. BMC Infectious Diseases. 2021 Mar 11;21(1):257.

47. Lauer SA, Grantz KH, Bi Q, Jones FK, Zheng Q, Meredith HR, et al. The Incubation Period of Coronavirus Disease 2019 (COVID-19) From Publicly Reported Confirmed Cases: Estimation and Application. Annals of Internal Medicine. 2020 Mar 10;172(9):577–82.

48. Fung HF, Martinez L, Alarid-Escudero F, Salomon JA, Studdert DM, Andrews JR, et al. The Household Secondary Attack Rate of Severe Acute Respiratory Syndrome Coronavirus 2 (SARS-CoV-2): A Rapid Review. Clinical Infectious Diseases. 2020 Oct 12;ciaa1558.

49. Madewell ZJ, Yang Y, Longini IM, Halloran ME, Dean NE. Household Transmission of SARS-CoV-2: A Systematic Review and Meta-analysis. JAMA Netw Open. 2020 Dec 14;3(12):e2031756.

50. Byambasuren O, Cardona M, Bell K, Clark J, McLaws M-L, Glasziou P. Estimating the extent of asymptomatic COVID-19 and its potential for community transmission: Systematic review and meta-analysis. Official Journal of the Association of Medical Microbiology and Infectious Disease Canada. 2020 Dec 1;5(4):223–34.

51. Han MS, Choi EH, Chang SH, Jin B-L, Lee EJ, Kim BN, et al. Clinical Characteristics and Viral RNA Detection in Children With Coronavirus Disease 2019 in the Republic of Korea. JAMA Pediatrics [Internet]. 2020 Aug 28 [cited 2020 Dec 12]; Available from: https://doi.org/10.1001/jamapediatrics.2020.3988

52. He X, Lau EHY, Wu P, Deng X, Wang J, Hao X, et al. Temporal dynamics in viral shedding and transmissibility of COVID-19. Nature Medicine. 2020 May;26(5):672–5.

53. Digest of Education Statistics, 2019 [Internet]. National Center for Education Statistics; [cited 2020 Dec 12]. Available from: https://nces.ed.gov/programs/digest/d19/tables/dt19_216.40.asp?current=yes

54. COVID-19 Projections Using Machine Learning [Internet]. COVID-19 Projections Using Machine Learning. [cited 2021 Apr 21]. Available from: https://covid19-projections.com/

55. Steel K, Davies B. COVID-19 Infection Survey: methods and further information [Internet]. 2021. Available from: https://www.ons.gov.uk/peoplepopulationandcommunity/healthandsocialcare/conditionsanddiseases/methodologies/covid19infectionsurveypilotmethodsandfurtherinformation

56. Waller A. About 80 Percent of K-12 Teachers and Staff Have Gotten a Covid-19 Vaccine Dose. The New York Times [Internet]. 2021 Apr 6 [cited 2021 May 12]; Available from: https://www.nytimes.com/live/2021/04/06/world/covid-vaccine-coronavirus-cases

57. Larremore DB, Wilder B, Lester E, Shehata S, Burke JM, Hay JA, et al. Test sensitivity is secondary to frequency and turnaround time for COVID-19 screening. Sci Adv [Internet]. 2021 Jan 1 [cited 2021 Mar 15];7(1). Available from: https://www.ncbi.nlm.nih.gov/pmc/articles/PMC7775777/

58. Atkeson A, Droste M, Mina MJ, Stock JH. Economic Benefits of COVID-19 Screening Tests with a Vaccine Rollout [Internet]. Health Economics; 2021 Mar [cited 2021 Mar 15]. Available from: http://medrxiv.org/lookup/doi/10.1101/2021.03.03.21252815

59. Cevik M, Tate M, Lloyd O, Maraolo AE, Schafers J, Ho A. SARS-CoV-2, SARS-CoV, and MERS-CoV viral load dynamics, duration of viral shedding, and infectiousness: a systematic review and meta-analysis. The Lancet Microbe. 2021 Jan 1;2(1):e13–22.

60. Wyllie AL, Fournier J, Casanovas-Massana A, Campbell M, Tokuyama M, Vijayakumar P, et al. Saliva or Nasopharyngeal Swab Specimens for Detection of SARS-CoV-2. New England Journal of Medicine [Internet]. 2020 Aug 28 [cited 2021 Apr 19]; Available from: https://www.nejm.org/doi/10.1056/NEJMc2016359

61. CDC. COVID-19 and Your Health [Internet]. Centers for Disease Control and Prevention. 2020 [cited 2021 Jan 4]. Available from: https://www.cdc.gov/coronavirus/2019-ncov/if-you-are-sick/quarantine.html

62. Pollock NR, Jacobs JR, Tran K, Cranston AE, Smith S, O’Kane CY, et al. Performance and Implementation Evaluation of the Abbott BinaxNOW Rapid Antigen Test in a High-Throughput Drive-Through Community Testing Site in Massachusetts. J Clin Microbiol. 2021 Apr 20;59(5):e00083–21.

63. Faherty LJ, Master BK, Steiner ED, Kaufman JH, Predmore Z, Stelitano L, et al. COVID-19 Testing in K–12 Schools: Insights from Early Adopters. 2021 Mar 9 [cited 2021 Aug 9]; Available from: https://www.rand.org/pubs/research_reports/RRA1103-1.html

64. Medicare Administrative Contractor (MAC) COVID-19 Test Pricing. :2.

65. Gewertz C. Testing for COVID-19 at School: Frequently Asked Questions. Education Week [Internet]. 2021 Mar 16 [cited 2021 Aug 9]; Available from: https://www.edweek.org/leadership/should-schools-test-students-and-staff-for-covid-19/2021/03

66. Paltiel AD, Zheng A, Sax PE. Clinical and Economic Effects of Widespread Rapid Testing to Decrease SARS-CoV-2 Transmission. Ann Intern Med. 2021 Jun 15;174(6):803–10.

67. Bastos ML, Perlman-Arrow S, Menzies D, Campbell JR. The Sensitivity and Costs of Testing for SARS-CoV-2 Infection With Saliva Versus Nasopharyngeal Swabs. Ann Intern Med. 2021 Jan 12;174(4):501–10.

68. Workman S, Jessen-Howard S. The True Cost of Providing Safe Child Care During the Coronavirus Pandemic [Internet]. Center for American Progress. [cited 2021 Apr 20]. Available from: https://www.americanprogress.org/issues/early-childhood/reports/2020/09/03/489900/true-cost-providing-safe-child-care-coronavirus-pandemic/

69. Parents and the High Cost of Child Care [Internet]. ChildCare Aware of America; 2017 [cited 2021 May 2]. Available from: https://www.childcareaware.org/wp-content/uploads/2017/12/2017_CCA_High_Cost_Report_FINAL.pdf

## References

1. Bilinski A, Salomon JA, Giardina J, Ciaranello A, Fitzpatrick MC. Passing the Test: A Model-Based Analysis of Safe School-Reopening Strategies. Ann Intern Med [Internet]. 2021 Jun 8 [cited 2021 Aug 9]; Available from: https://www.acpjournals.org/doi/10.7326/M21-0600

2. Gatto M, Bertuzzo E, Mari L, Miccoli S, Carraro L, Casagrandi R, et al. Spread and dynamics of the COVID-19 epidemic in Italy: Effects of emergency containment measures. Proc Natl Acad Sci USA. 2020 May 12;117(19):10484–91.

3. He X, Lau EHY, Wu P, Deng X, Wang J, Hao X, et al. Temporal dynamics in viral shedding and transmissibility of COVID-19. Nat Med. 2020 Apr 15;1–4.

4. Kerr CC, Stuart RM, Mistry D, Abeysuriya RG, Hart G, Rosenfeld K, et al. Covasim: an agent-based model of COVID-19 dynamics and interventions. medRxiv. 2020 May 15;2020.05.10.20097469.

5. He D, Zhao S, Lin Q, Zhuang Z, Cao P, Wang MH, et al. The relative transmissibility of asymptomatic COVID-19 infections among close contacts. International Journal of Infectious Diseases. 2020 May 1;94:145–7.

6. Li Q, Guan X, Wu P, Wang X, Zhou L, Tong Y, et al. Early Transmission Dynamics in Wuhan, China, of Novel Coronavirus–Infected Pneumonia. New England Journal of Medicine. 2020 Jan 29;0(0):null.

7. Firth JA, Hellewell J, Klepac P, Kissler SM, CMMID COVID-19 working group, Kucharski AJ, et al. Combining fine-scale social contact data with epidemic modelling reveals interactions between contact tracing, quarantine, testing and physical distancing for controlling COVID-19 [Internet]. Epidemiology; 2020 May [cited 2020 Jun 2]. Available from: http://medrxiv.org/lookup/doi/10.1101/2020.05.26.20113720

8. Endo A, Centre for the Mathematical Modelling of Infectious Diseases COVID-19 Working Group, Abbott S, Kucharski AJ, Funk S. Estimating the overdispersion in COVID-19 transmission using outbreak sizes outside China. Wellcome Open Res. 2020 Apr 9;5:67.

9. Byambasuren O, Cardona M, Bell K, Clark J, McLaws M-L, Glasziou P. Estimating the extent of asymptomatic COVID-19 and its potential for community transmission: Systematic review and meta-analysis. Official Journal of the Association of Medical Microbiology and Infectious Disease Canada. 2020 Dec 1;5(4):223–34.

10. Fontanet A, Grant R, Tondeur L, Madec Y, Grzelak L, Cailleau I, et al. SARS-CoV-2 infection in primary schools in northern France: A retrospective cohort study in an area of high transmission. medRxiv. 2020 Jun 29;2020.06.25.20140178.

11. Stein-Zamir C, Abramson N, Shoob H, Libal E, Bitan M, Cardash T, et al. A large COVID-19 outbreak in a high school 10 days after schools’ reopening, Israel, May 2020. Eurosurveillance. 2020 Jul 23;25(29):2001352.

12. Dattner I, Goldberg Y, Katriel G, Yaari R, Gal N, Miron Y, et al. The role of children in the spread of COVID-19: Using household data from Bnei Brak, Israel, to estimate the relative susceptibility and infectivity of children [Internet]. Infectious Diseases (except HIV/AIDS); 2020 Jun [cited 2020 Jun 19]. Available from: http://medrxiv.org/lookup/doi/10.1101/2020.06.03.20121145

13. Park YJ, Choe YJ, Park O, Park SY, Kim Y-M, Kim J, et al. Early Release - Contact Tracing during Coronavirus Disease Outbreak, South Korea, 2020 - Volume 26, Number 10— October 2020 - Emerging Infectious Diseases journal - CDC. [cited 2020 Aug 1]; Available from: https://www.nc.cdc.gov/eid/article/26/10/20-1315_article

14. Fontanet A, Tondeur L, Madec Y, Grant R, Besombes C, Jolly N, et al. Cluster of COVID-19 in northern France: A retrospective closed cohort study. medRxiv. 2020 Apr 23;2020.04.18.20071134.

15. Mutesa L, Ndishimye P, Butera Y, Souopgui J, Uwineza A, Rutayisire R, et al. A pooled testing strategy for identifying SARS-CoV-2 at low prevalence. Nature. 2021 Jan;589(7841):276–80.

16. Lohse S, Pfuhl T, Berkó-Göttel B, Rissland J, Geißler T, Gärtner B, et al. Pooling of samples for testing for SARS-CoV-2 in asymptomatic people. The Lancet Infectious Diseases. 2020 Nov 1;20(11):1231–2.

17. Goldstein E, Lipsitch M, Cevik M. On the Effect of Age on the Transmission of SARS-CoV-2 in Households, Schools, and the Community. The Journal of Infectious Diseases [Internet]. 2020 Oct 29 [cited 2021 Jan 24];(jiaa691). Available from: https://doi.org/10.1093/infdis/jiaa691

18. Madewell ZJ, Yang Y, Longini IM, Halloran ME, Dean NE. Household Transmission of SARS-CoV-2: A Systematic Review and Meta-analysis. JAMA Netw Open. 2020 Dec 14;3(12):e2031756.

19. Fung HF, Martinez L, Alarid-Escudero F, Salomon JA, Studdert DM, Andrews JR, et al. The Household Secondary Attack Rate of Severe Acute Respiratory Syndrome Coronavirus 2 (SARS-CoV-2): A Rapid Review. Clinical Infectious Diseases. 2020 Oct 12;ciaa1558.

20. Alonso S, Alvarez-Lacalle E, Català M, López D, Jordan I, García-García JJ, et al. Age-dependency of the Propagation Rate of Coronavirus Disease 2019 Inside School Bubble Groups in Catalonia, Spain. The Pediatric Infectious Disease Journal [Internet]. 2021 Jul 29 [cited 2021 Aug 9]; Available from: https://journals.lww.com/pidj/Abstract/9000/Age_dependency_of_the_Propagation_Rate _of.95714.aspx

21. Doyle T. COVID-19 in Primary and Secondary School Settings During the First Semester of School Reopening — Florida, August–December 2020. MMWR Morb Mortal Wkly Rep [Internet]. 2021 [cited 2021 May 14];70. Available from: https://www.cdc.gov/mmwr/volumes/70/wr/mm7012e2.htm

22. Gettings J. Mask Use and Ventilation Improvements to Reduce COVID-19 Incidence in Elementary Schools — Georgia, November 16–December 11, 2020. MMWR Morb Mortal Wkly Rep [Internet]. 2021 [cited 2021 Aug 9];70. Available from: https://www.cdc.gov/mmwr/volumes/70/wr/mm7021e1.htm

23. Falk A. COVID-19 Cases and Transmission in 17 K–12 Schools — Wood County, Wisconsin, August 31–November 29, 2020. MMWR Morb Mortal Wkly Rep [Internet]. 2021 [cited 2021 Apr 21];70. Available from: https://www.cdc.gov/mmwr/volumes/70/wr/mm7004e3.htm

24. Lessler J, Grabowski MK, Grantz KH, Badillo-Goicoechea E, Metcalf CJE, Lupton-Smith C, et al. Household COVID-19 risk and in-person schooling. Science [Internet]. 2021 Apr 29 [cited 2021 May 21]; Available from: https://science.sciencemag.org/content/early/2021/04/28/science.abh2939

25. The Association of Opening K-12 Schools with the Spread of COVID-19 in the United States: County-Level Panel Data Analysis [Internet]. [cited 2021 Aug 9]. Available from: https://www.medrxiv.org/content/10.1101/2021.02.20.21252131v2

26. Goldhaber D, Imberman S, Strunk KO, Hopkins B, Brown N, Harbatkin E, et al. To What Extent Does In-Person Schooling Contribute to the Spread of COVID-19? Evidence from Michigan and Washington [Internet]. 2020 Dec [cited 2021 Jan 24]. Available from: https://epicedpolicy.org/does-in-person-schooling-contribute-to-the-spread-of-covid-19/

27. CDC. Delta Variant: What We Know About the Science [Internet]. Centers for Disease Control and Prevention. 2021 [cited 2021 Aug 9]. Available from: https://www.cdc.gov/coronavirus/2019-ncov/variants/delta-variant.html

28. Campbell F, Archer B, Laurenson-Schafer H, Jinnai Y, Konings F, Batra N, et al. Increased transmissibility and global spread of SARS-CoV-2 variants of concern as at June 2021. Eurosurveillance. 2021 Jun 17;26(24):2100509.

29. Sheikh A, McMenamin J, Taylor B, Robertson C, Public Health Scotland and the EAVE II Collaborators. SARS-CoV-2 Delta VOC in Scotland: demographics, risk of hospital admission, and vaccine effectiveness. Lancet. 2021 Jun 26;397(10293):2461–2.

30. Lopez Bernal J, Andrews N, Gower C, Gallagher E, Simmons R, Thelwall S, et al. Effectiveness of Covid-19 Vaccines against the B.1.617.2 (Delta) Variant. New England Journal of Medicine. 2021 Jul 21.

